# Modelling the effect of long-lasting insecticidal nets on malaria transmission dynamics in Kebbi State, Nigeria

**DOI:** 10.64898/2026.01.15.26344194

**Authors:** Emmanuel A. Bakare, Idowu I. Olasupo, Micheal Imoudu, Afeez Abidemi, Deborah O. Daniel, Samuel A. Osikoya, Oluwaseun A. Mogbojuri, Aaron O. Nwana, Dolapo O. Oniyelu, Ronke D. Olorunfemi, Samson O. Olagbami, Dorcas O. Agboola, Steven I. Ikediashi, Ayomide Adedeji, Oluwaponmile Ikuseka, Micheal Fadairo, Odunayo Odewale, Goodness Onwuka, Aurel Hansinon, Dolapo A. Bakare, Lisa J. White, Nakul Chitnis, Olusola Oresanya, Chukwu Okoronkwo, Eze Nelson, Segun Kosoko, Gabriel I. Ogban

## Abstract

**Background/Aim:** Malaria, in Nigeria, is a disease of public health concern that has caused both morbidity and mortality, with the highest prevalence in Kebbi State. Long-lasting insecticidal nets (LLINs) have been instrumental in controlling the burden of the disease. This study aims to assess the effect of LLINs on malaria transmission dynamics in Kebbi State, Nigeria.

**Methods:** Routine data for the confirmed uncomplicated malaria cases in Kebbi State, Nigeria, from January 2015 to May 2024 were used to understand the transmission dynamics within the population. A deterministic model was developed to capture the malaria transmission dynamics in Kebbi State. Qualitative analysis was carried out, establishing the positivity and boundedness of solutions to ensure the biological feasibility of the model. The disease-free equilibrium was analyzed for stability, revealing under which condition the disease is eradicated. Effective reproduction number, ℛ_*e*_, was derived, governing the disease persistent in the presence of intervention. The endemic equilibrium was further examined to indicate situations where the disease persist in the population. The model was fitted to Kebbi state monthly malaria cases using the least squares estimation method implemented in R. Numerical simulations were performed using the R software. Prediction scenarios of malaria cases considering different usage levels of LLINs (38.2%, 50% and 80%) are visualised through the simulation of the malaria model.

**Results:** The result showed that if there had been 80% sustained level LLINs usage as proposed by NMEP since 2015, there would have been about 5.2 million malaria cases averted, which corresponds to 97.98% reduction. However, moving forward, if 80% usage can be achieved and sustained, about 3 million malaria cases would be averted by May 2029, signifying an impressive reduction of 78.93% in incidence.

**Conclusion:** We conclude that in order to significantly reduce malaria incidence in Kebbi State to its bearest minimum, health policymakers and decision makers in Nigeria should prioritise scaling up LLINs usage in the State by expanding LLINs distribution and improving malaria education in the .

## 1 Introduction

Malaria is a disease caused by the parasite of the genus *Plasmodium* transmitted through the bite of infected female Anopheles mosquitoes [1, 2]. Five species of *Plasmodium* are known to cause malaria in humans -*Plasmodium falciparum* (*P. falciparum*), *P. vivax, P. ovale, P. malariae*, and the simian species *P. knowlesi* [1]. Two species that pose the greatest threat are *P. falciparum* and *P. vivax* [1]. *P. falciparum* is the most virulent species, accounting for over 97% of the total malaria cases and is the predominant species in Nigeria [1, 3].

Malaria is a disease of public health concern that has caused both morbidity and mortality [1]. It remains a global health issue, with sub-Saharan Africa being the most affected region, accounting for over 90% of malaria cases and deaths [4]. According to the World Health Organization (WHO), 249 million malaria cases were reported globally in 2022, resulting in approximately 608,000 deaths [4, 5, 6]. The African continent bears a great share of this burden, with Nigeria accounting for 27% of the global malaria burden with an estimated 68 million cases and 194,000 deaths due to the disease in 2021 [7]. Also, of all the 36 States in Nigeria and the Federal Capital Territory (FCT), Kebbi State records the highest malaria prevalence with microscopy of 49% [6, 7].

In the year 2001, Nigeria launched the National Malaria Control Program (NMCP), later renamed the National Malaria Elimination Program (NMEP), to address the burden of malaria in the country [8]. This was to guide the nation to achieve malaria pre-elimination and elimination as proposed in her national malaria elimination strategic plan (NMSP) [9]. Thus far, the country has successfully implemented four of her strategic plans and is currently in the fifth plan, which will elapse by the end of 2025 [8, 9]. Currently, major interventions such as long-lasting insecticidal nets (LLINs), case management, intermittent preventive treatment (IPTp), and seasonal malaria chemoprevention (SMC) have been deployed in Nigeria [8, 9]. NMEP proposes 80% scale-up of LLINs distributions in the country at 3-year intervals, owing to its proven efficacy and cost-effectiveness in reducing malaria morbidity and mortality across various regions [10]. However, this proposed plan has not been sustained over the years, which makes malaria remain highly endemic in the country, particularly in Kebbi State with the highest prevalence [11].

Malaria transmission in Kebbi State is seasonal, with peaks occurring during the rainy season, which usually spans from June to October [6, 12]. This period provides ideal conditions for the breeding of Anopheles mosquitoes, the primary vectors of malaria [6, 12]. Thus far, there have been three rollouts of LLINs campaigns in the State in 2009, 2014, and 2022 [6]. An estimate of 3.5 million LLINs have been distributed through mass distribution campaigns [6, 8]. The number of people who slept under LLINs the night before the Demographic and Health Surveys (DHS)/MIS surveys increased from 37.6% in 2015 to 38.2% in 2021 [6, 8]. The widespread deployment in endemic regions, particularly in Nigeria, has been instrumental in reducing the incidence and prevalence of the disease [13].

Among the various aforementioned control measures, LLINs have been a major intervention in the fight against malaria in Nigeria, particularly Kebbi State [6, 8]. They represent a significant advancement over earlier mosquito nets, as they incorporate insecticides that repel or kill mosquitoes upon contact, thereby reducing mosquito survival and preventing bites [14]. Unlike traditional insecticide-treated nets (ITNs), LLINs maintain effective insecticide levels for at least three years, even after multiple washes [13, 14]. Nevertheless, the efficacy of LLINs is often compromised by improper use. Discomfort and misconceptions about LLINs contribute to low utilization rates in some communities [3]. Furthermore, the growing prevalence of insecticide resistance among mosquito populations poses a substantial challenge to the sustainability of LLINs as a control measure [15]. This intervention has been reported to be particularly effective in high-transmission regions in Nigeria [11].

Studies have highlighted the importance of LLIN as an important and mainstay intervention against malaria in Nigeria. The work of Saleh et al. [16] revealed that there is a positive correlation between the use of LLINs and a decline in malaria prevalence from the 2015 MIS from across the six geopolitical zones in Nigeria [6, 16]. Hence, there has been a recommendation to increase the distribution and access to LLINs to individuals [16]. The study’s outcome again underscores LLIN as an important intervention tool against malaria and its unwanted consequences.

Over the years, mathematical modelling has proven to be an essential tool for understanding disease transmission dynamics and informing intervention strategies [17, 18]. For instance, Bakare et al. (2021) [19] developed a mathematical model to analyse the potential impact of multiple control interventions such as educational campaigns, insecticide-treated nets, indoor residual spraying (IRS), destruction of mosquito breeding sites, treatment with artemisinin-based combination treatment (ACT) on malaria transmission in limited resources community. Gimba and Bala (2017) [20] investigated the effect of bed net use, insecticide-treated nets (ITNs), temperature, and treatment on malaria transmission. The study’s results showed that the mosquito biting rate peak occurs at a certain range of temperature. Also, the research findings showed that a combination of treatment and ITN’s usage is the most effective intervention strategy. The study highlights that mosquito biting and death rates are the most important parameters driving malaria transmission, which suggests the effectiveness of LLINS. These results underscore the effectiveness of LLINs in reducing biting rates and increasing mosquito death rates due to the chemicals of the LLINs. Mukhtar et al. (2018) [21] developed a deterministic model that captures the effect of LLINs on malaria transmission in South Sudan. The study suggests that the effective use of LLINs can reduce ℛ_*o*_ and malaria transmission. It also advocates for scaling up LLIN distribution, particularly targeting households in high-risk malaria areas. Ngonghala et al. (2014) [22] developed a mathematical model that represents the decrease in the effectiveness of LLINs due to physical and chemical decay and human behaviour on malaria transmission. Another work of Ngonghala (2022) [23] studied the effects of ITNs, decay in ITNs efficacy over time, and mosquito-resistant insecticide on malaria transmission dynamics accounting for asymptomatic individuals. The findings highlight the importance of replacing ITNs before their prescribed lifespans or designing longer-lasting ITNs. Additionally, piperonyl butoxide (PBO) ITNs, which counteract insecticide resistance, are shown to be more effective than regular ITNs in controlling malaria.

Despite the reports on the effectiveness of LLINs in reducing malaria transmission, significant gaps remain in ensuring their sustained impact, particularly in regions with high malaria prevalence like Kebbi State. Challenges such as maintaining high LLIN coverage and usage, addressing insecticide resistance, and optimizing distribution strategies hinder long-term malaria control efforts. Moreover, existing studies often focus on national or sub-regional malaria trends without detailed studies on the effect of LLINs at the State level, particularly in Kebbi State. This study aims to assess the effect of LLINs on malaria transmission dynamics in Kebbi State by considering varying LLINs usage scenarios and also quantifying the number of cases averted with LLINs. This work seeks to provide evidence-based recommendations for improving malaria elimination efforts in the State.

The remaining part of the article is organized as follow. Section 2 details the materials and methods, which includes the study area, data description, model formulation methods, qualitative analysis of the model, and model calibration. Section 3 presents the results, and section 4 presents the discussion of the results. Section 5 summarizes the key findings, limitations, and recommendations.

## 2 Materials and methods

### 2.1 Study area

Kebbi State is a State in northwest Nigeria that lies between Zamfara and Niger State in the south and between Sokoto to the west [24, 25]. The total land area of Kebbi State is around 36,800 square kilometers [26]. There are two primary seasons in the State: the Rainy Season (May to October), which has the highest rainfall from July to September, and the Dry Season (November to April), which is characterized by decreased humidity and dry winds, temperatures, particularly in April, is often the warmest month [27].

### 2.2 Data description

Routine data for Kebbi State from January 2015 - May 2024 was obtained from National Malaria Data Repository (NMDR) of NMEP [28]. The data are not publicly accessible due to governmental policies to maintain confidentiality requirements, but access can be granted upon request following necessary approvals. Variables present in the data comprises of 113 observations and includes: confirmed uncomplicated malaria, persons with clinically diagnosed malaria, children under 5 years who received LLIN, persons with confirmed uncomplicated malaria treated with ACT, persons clinically diagnosed with malaria treated with ACT, e.t.c. The confirmed uncomplicated malaria cases in the State will be used to understand the transmission dynamics within the population to analyse the transmission dynamics of malaria in the State. The time series plot of the confirmed uncomplicated malaria case incidence trend is shown in Figure 1b.

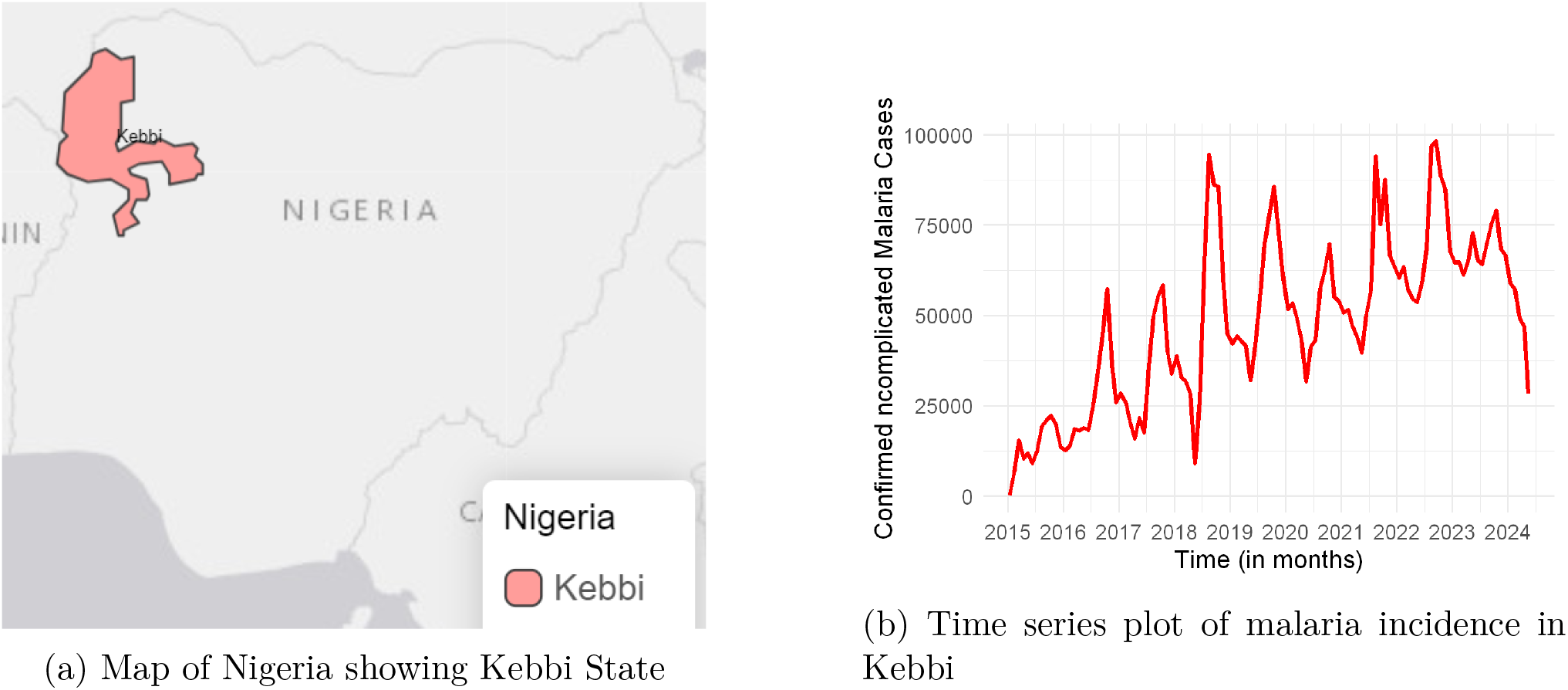

### 2.3 Model formulation

We develop a deterministic compartmental model that incorporates the effect of LLIN. The model is divided into two major groups: the human population and the female *Anopheles* mosquito population. The human population is subdivided into four compartments, namely, susceptible human *S*_*h*_(*t*), pre-infectious human *E*_*h*_(*t*), symptomatic infectious human *I*_*h*_(*t*), and asymptomatic infectious human *A*_*h*_(*t*). Thus, the total human population *N*_*h*_(*t*), is given as

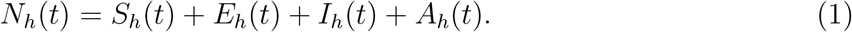

In our model, it is assumed that after recovery, individuals return to being susceptible. This assumption is a departure from what is seen in many malaria models in literature that move people to recovered class after recovery and thereafter to susceptible class. The latter approach has an underlying assumption that recovered individuals have anti-parasite immunity during the period they are in the recovered class, that is, individuals in the recovered class are assumed to be immune to infection, which is against the reality of *Plasmodium falciparum* malaria [29]. We acknowledge that recovery from *Plasmodium falciparum* malaria confers anti-disease immunity, that is, recovered individuals are only immune to malaria symptoms rather than the infection for some time [29]. This gives us an understanding that an individual can still be infected if exposed to infectious mosquito bites after recovery. It is known that even the anti-disease immunity is not so certain, since there is high variability in the level of this immunity among individuals, hence to simplify the complexity that may arise due to this variability, we assume that after recovery, individuals return to susceptible class.

The mosquito population is divided into three compartments; susceptible mosquitoes *S*_*m*_(*t*), pre-infectious mosquitoes *E*_*m*_(*t*), and infectious mosquitoes *I*_*m*_(*t*). The total mosquito population *N*_*m*_(*t*), is thus given as:

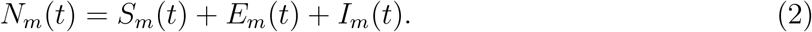

We assume a constant recruitment rate Λ_*h*_ into the susceptible human population. The susceptible human population is further increased when symptomatic infectious humans recover naturally or due to treatment at rate *θ*_*I*_ + *u*_2_ and become susceptible again. The population decreases when susceptible humans get infected at a rate 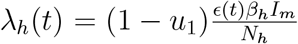, where *β*_*h*_ is the probability of transmission of malaria infection from an infectious mosquito to a susceptible human and (1 − *u*_1_) represents the reduction in transmission due to LLIN, with *u*_1_, (0 ≤ *u*_1_ < 1) being the protective effectiveness of LLIN.

Adapting the forcing function employed in [30], we set the time-dependent function

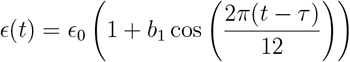

to be the seasonally-forced mosquito biting rate which accounts for the seasonal patterns observed in the data, where *ϵ*_0_ is the per capita (baseline) biting rate of mosquitoes, *b*_1_ is the amplitude and *τ* is the phase shift. The susceptible human population further decreases due to natural death at rate *µ*_*h*_.

Thus, the rate of change of the susceptible human population at any given time *t* is given as

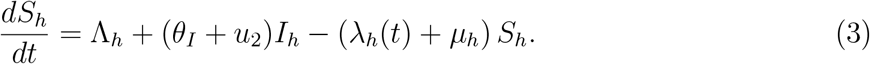

Infected humans transit to the pre-infectious class at a rate 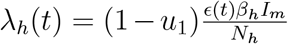. At the end of the latent phase, individuals leave the pre-infectious class and become infectious at the rate *σ*_*h*_. A further decrease occur in the pre-infectious human class via natural death at the rate *µ*_*h*_. So, the rate of change of the pre-infectious human population is given as

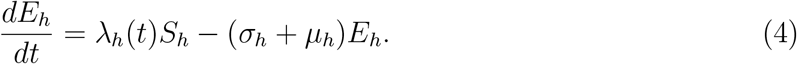

The symptomatic infectious compartment is increased when a proportion *ρ* transit from the pre-infectious class and become symptomatic at rate *σ*_*h*_. Also, the population of symptomatic infectious human is increased when asymptomatic individuals get re-infected and become symptomatic at rate *ηλ*_*h*_(*t*). The symptomatic infectious class is reduced either via natural recovery or treatment-induced recovery at rates *θ*_*I*_ or *u*_2_ respectively and join the susceptible class. Further reduction is due to natural death at rate *µ*_*h*_ and malaria-induced death at rate *δ*. Consequently, the rate of change of the symptomatic infectious population is given as

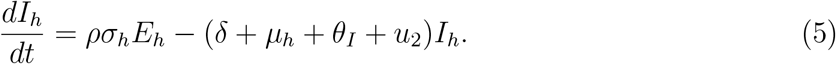

Similarly, the asymptomatic compartment is increased when a proportion (1 − *ρ*) moves from the exposed class and become asymptomatic at the rate *σ*_*h*_. We assume that there is no explicit recovery pathway for the asymptomatic infectious individuals. This is in line with the dynamics of malaria in endemic settings such as Kebbi State, where asymptomatic individuals in the population are constantly exposed to infectious bites, and suffer re-infection, leading to spike in parasite load and development of symptoms [31, 32, 33]. Thus, the asymptomatic infectious population is depleted when asymptomatic individuals get re-infected and become symptomatic at rate *ηλ*_*h*_(*t*), and via natural death at rate *µ*_*h*_. Thus, the rate of change of the asymptomatic infectious population at time *t* is given as

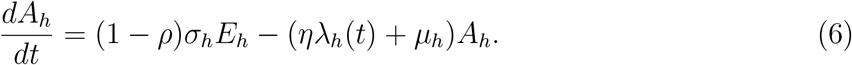

Female Anopheles mosquitoes are recruited into the susceptible population at rate Λ_*m*_. The population is reduced when susceptible female Anopheles mosquitoes get infected at a rate 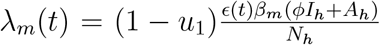 where, *β*_*m*_ is the probability of transmission from an infectious human to a susceptible mosquito and *ϕ* (0 < *ϕ* < 1) is a modification (reduction) parameter due to reduced infectivity of symptomatic individuals compared to asymptomatic individuals. The population is further reduced due to natural death at rate *µ*_*m*_. Thus, the rate of change of the susceptible mosquito population is given as

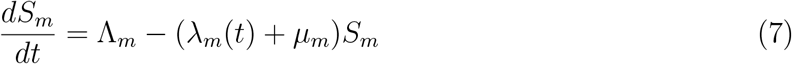

Newly infected female Anopheles mosquitoes move to the pre-infectious class at a rate 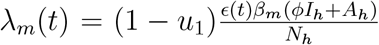. The population of pre-infectious mosquitoes decreases as they either become infectious, transitioning to the infectious class at a rate *σ*_*m*_, or die naturally at a rate *µ*_*m*_. So, the rate of change of the population of pre-infectious mosquitoes is given as

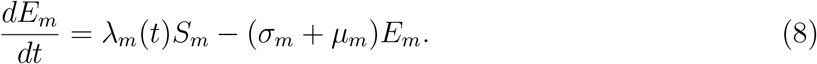

The infectious mosquito compartment is generated as a result of progression from the preinfectious compartment at a rate *σ*_*m*_. The population of infectious mosquitoes is decreased due to natural death at a rate *µ*_*m*_. Thus, the rate of change of the population of infectious mosquito is obtained as

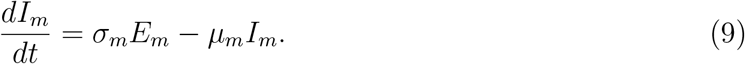

From the discussion above, we obtain a seven-dimensional system of nonlinear differential equations given in equation (10). The State variables are described in Table 1 while the description of the model parameters are presented in Table 2. Figure 2 illustrates the transmission dynamics of malaria between human and mosquito populations incorporating LLIN intervention.

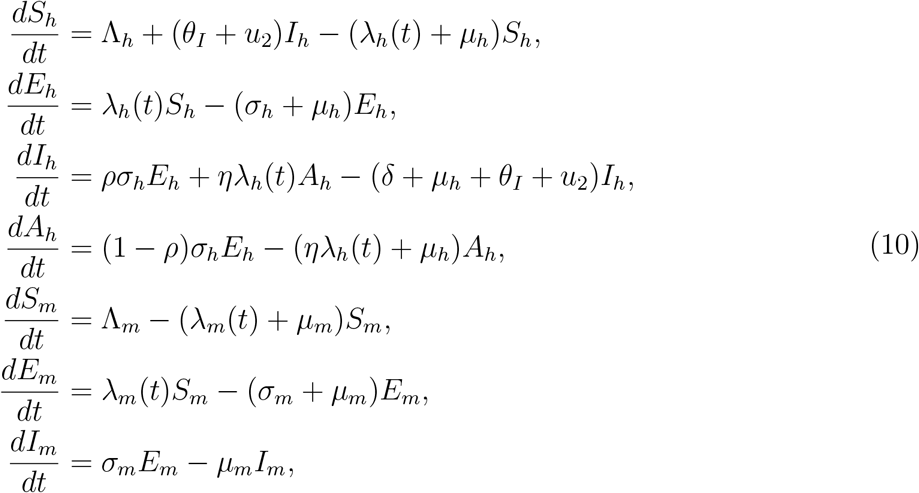

with the following initial conditions:

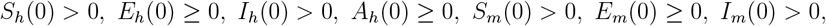

where

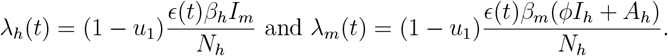

**Table 1:** Description of variables of system (10)

| Variable | Description |
| --- | --- |
| $S_h$ | Population of susceptible individuals |
| $E_h$ | Population of pre-infectious individuals |
| $I_h$ | Population of symptomatic infectious individuals |
| $A_h$ | Population of asymptomatic individuals |
| $S_m$ | Population of susceptible female Anopheles mosquitoes |
| $E_m$ | Population of pre-infectious female Anopheles mosquitoes |
| $I_m$ | Population of infectious female Anopheles mosquitoes |

**Table 2:** Description of model parameters.

| Parameter | Description |
| --- | --- |
| $\Lambda_h$ | Recruitment rate of humans |
| $\Lambda_m$ | Recruitment rate of mosquitoes |
| $\beta_h$ | Transmission probability from an infectious mosquito to a susceptible human |
| $\beta_m$ | Transmission probability from an infectious human to a susceptible mosquito |
| $\epsilon_0$ | Per capita (baseline) biting rate of mosquitoes |
| $\rho$ | Proportion of exposed individuals progressing to the symptomatic infectious class |
| $1 - \rho$ | Proportion of exposed individuals progressing to the asymptomatic infectious class |
| $\mu_h$ | Per capita natural death rate of humans |
| $\mu_m$ | Per capita natural death rate of mosquitoes |
| $\sigma_h$ | Progression rate exposed human class to infectious human class |
| $\sigma_m$ | Progression rate from exposed mosquito class to the infectious mosquito class |
| $\eta$ | Probability of asymptomatic infectious human getting more infectious bites |
| $\phi$ | Modification parameter |
| $\delta_h$ | Per capita disease-induced death rate for humans. |
| $\theta_I$ | Natural recovery rate of symptomatic individuals |
| $u_1$ | Protective effectiveness of LLINs |
| $u_2$ | Treatment rate |
| $b_1$ | Amplitude |
| $\tau$ | Phase shift |

**Figure 2.**
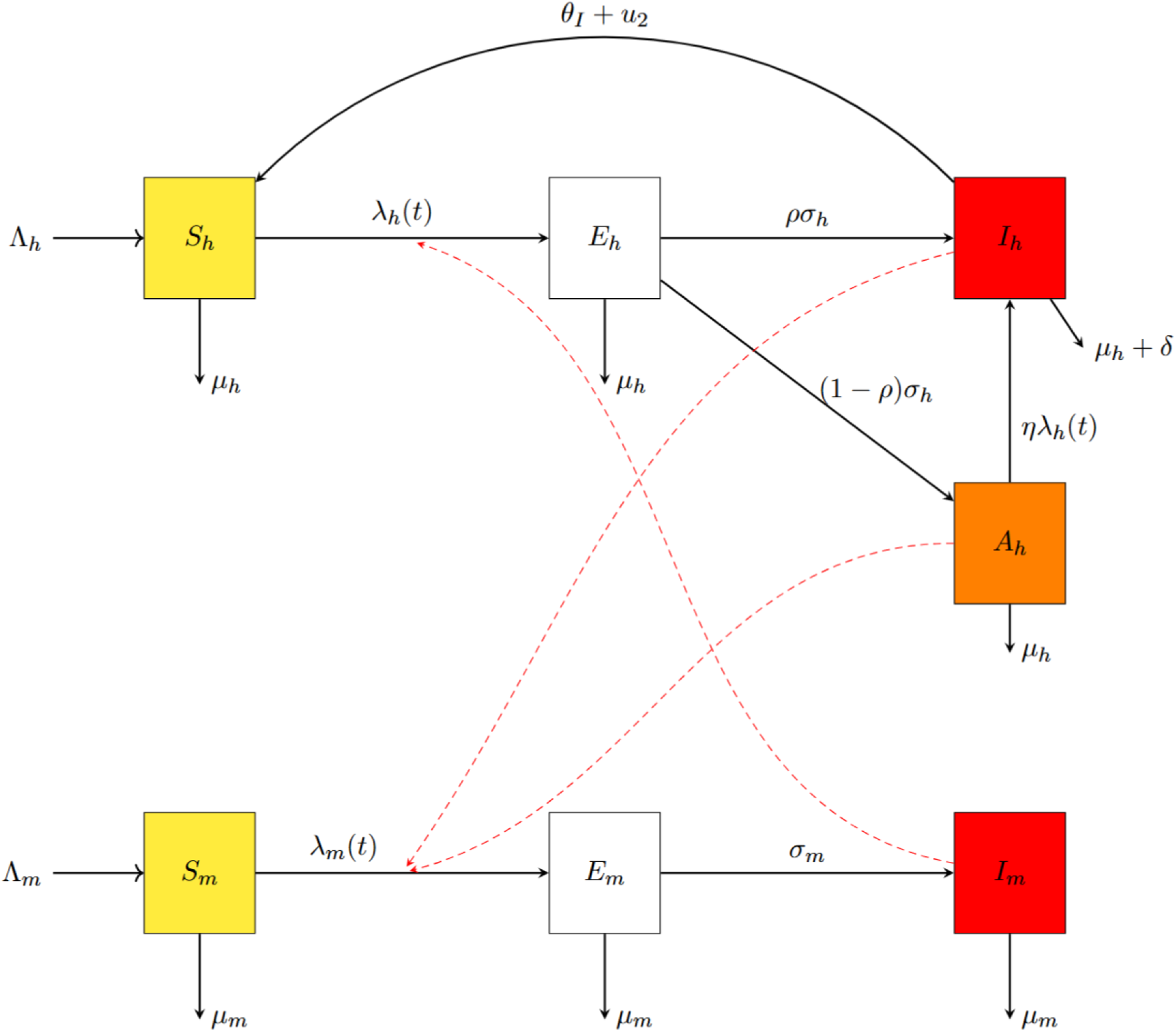
The flowchart of malaria transmission dynamics between the human and mosquito population (10)

### 2.4 Model calibration

This section presents the calibration process for system (10). Kebbi State monthly malaria cases from 2015 to 2024 were used to calibrate model (10) using the method of least squares estimate with R software. The method seeks to find the model parameter values that minimise the sum of squared errors between the model and the data, that is

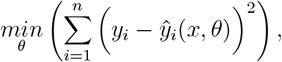

where *y*_*i*_ is the number of malaria cases at the *i*th month and *ŷ*_*i*_(*x, θ*) is the estimated malaria cases at the *i*th month which depend on the model State variable *x* and parameter *θ*.

The initial conditions for malaria model (10) are obtained as follows: The symptomatic infectious human population is taken to be the number of cases in January 2015, hence *I*_*h*_(0) = 177. We assume that the exposed human population *E*_*h*_(0) = 0. Asymptomatic infectious humans is assumed to be 5 times as much as *I*_*h*_(0), that is *A*_*h*_(0) = 5 × *I*_*h*_(0) = 885. The total human population of Kebbi State in 2015 was 4, 377, 770, so that the initial total human population is taken as *N*_*h*_(0) = 4, 377, 770. It follows that the initial susceptible human population is obtained as *S*_*h*_(0) = *N*_*h*_(0) − (*E*_*h*_(0) + *I*_*h*_(0) + *A*_*h*_(0)) = 4, 377, 770 − (177 + 885) = 4, 376, 708.

For the mosquito population, we assume that the initial total mosquito population is 5 times the total human population, that is, *N*_*m*_(0) = 5 × *N*_*h*_(0) = 21, 888, 850. The initial size of the exposed mosquito population is assumed to be *E*_*m*_(0) = 0, while the initial infected mosquito population is taken as *I*_*m*_(0) = 200, 000. Consequently, the initial susceptible mosquito population is obtained as *S*_*m*_(0) = 5 × 4, 377, 770 − 200, 000 = 21, 688, 850.

The model’s parameters are estimated via the calibration process except for the recruitment rate of humans Λ_*h*_, the recruitment rate of mosquito Λ_*m*_, natural death rate of humans and mosquitoes *µ*_*h*_ and *µ*_*m*_ respectively, and recovery rate for symptomatic humans *θ*_*I*_. The parameters *µ*_*h*_ and *µ*_*m*_ are calculated as the inverse of the average human lifespan in Kebbi State and life span of mosquito respectively, so that 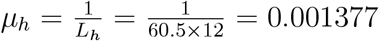 per month, where *L*_*h*_ = 60.5 × 12 represents the average human lifespan in Kebbi State (in months) [34]. and 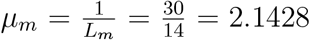 per month, where *L*_*m*_ = 14/30 represents the average lifespan of a mosquito (in months) [35]. The parameters Λ_*h*_ and Λ_*m*_ are calculated as follows: since the total population of Kebbi State is 4, 377, 770 in 2015, then, 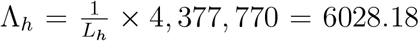, and 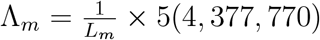. The recovery rate for symptomatic individuals is calculated by taking the inverse of its average infectious period, 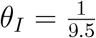 per month [36]. The progression rate of the exposed human class to the infectious human class and from the exposed mosquito class to the infectious mosquito class (*σ*_*h*_ and *σ*_*m*_) is obtained by the inverse of the average incubation period of *P. falciparum* parasite in humans and mosquitoes, 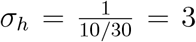 per month, 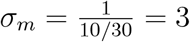 per month [36]. Protective effectiveness of LLINs, *u*_1_, is considered as a product of LLIN efficacy and LLIN usage. According to [37, 38], a newly acquired LLIN is expected to have an efficacy between 0.9 and 1. Thus, we set the LLIN efficacy as 0.9 in this study. The LLIN usage in 2015 is 0.382 [6]. Consequently, protective effectiveness of LLINs is calculated as *u*_1_ = 0.9 × 0.382 = 0.3438. In addition, the efficacy of ACT is reported to be 0.98 [6]. The treatment coverage (that is, proportion of confirmed uncomplicated malaria cases treated with ACT) is obtained from the data as 0.99. Based on therapeutic efficacy studies [39], it is expected that gametocyte clearance occurs within 3 to 28 days of ACT use. The treatment rate due to ACT, *u*_2_, is therefore obtained using the survival function to convert from proportion to rate as 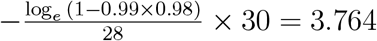 per month.

The remaining parameters *β*_*h*_, *β*_*m*_, *ρ, η, τ* and *b*_1_ are obtained using the least-square curve fitting method implemented in R programming. The result of fitting the model (10) with the actual Kebbi State malaria incidence data is displayed in Figure 3, while Table 3 gives the summary of the model parameter values obtained from the fitting process.

**Table 3:** Parameter values for the malaria transmission model.

| Parameter | Value | Unit | Source |
| --- | --- | --- | --- |
| $\beta_h$ | 0.13044 | Dimensionless | Fitted |
| $\beta_m$ | 0.01000 | Dimensionless | Fitted |
| $\mu_h$ | 0.00137 | Per month | [34] |
| $\mu_m$ | 2.1428 | Per month | [36] |
| $\rho$ | $\frac{1}{6}$ | Dimensionless | Fitted |
| $\sigma_h$ | 3 | Per month | [36] |
| $\sigma_m$ | 3 | Per month | [36] |
| $\theta_I$ | $\frac{1}{9.5}$ | Per month | [36] |
| $\eta$ | 0.07973 | Per month | Fitted |
| $\delta$ | $2.7 \times 10^{-3}$ | Per month | [36] |
| $\Lambda_h$ | 6028.18 | Per human per month | Derived |
| $\Lambda_m$ | 45966585 | Per mosquito per month | Derived |
| $\phi$ | 0.7 | Dimensionless | Assumed |
| $u_1$ | 0.3438 | Dimensionless | [6] |
| $u_2$ | 3.764 | Dimensionless | [36] |
| $\tau$ | 8.68604 | Dimensionless | Fitted |
| $b_1$ | 0.16400 | Dimensionless | Fitted |
| $\epsilon_0$ | 13 | Per month | [36] |

**Figure 3.**
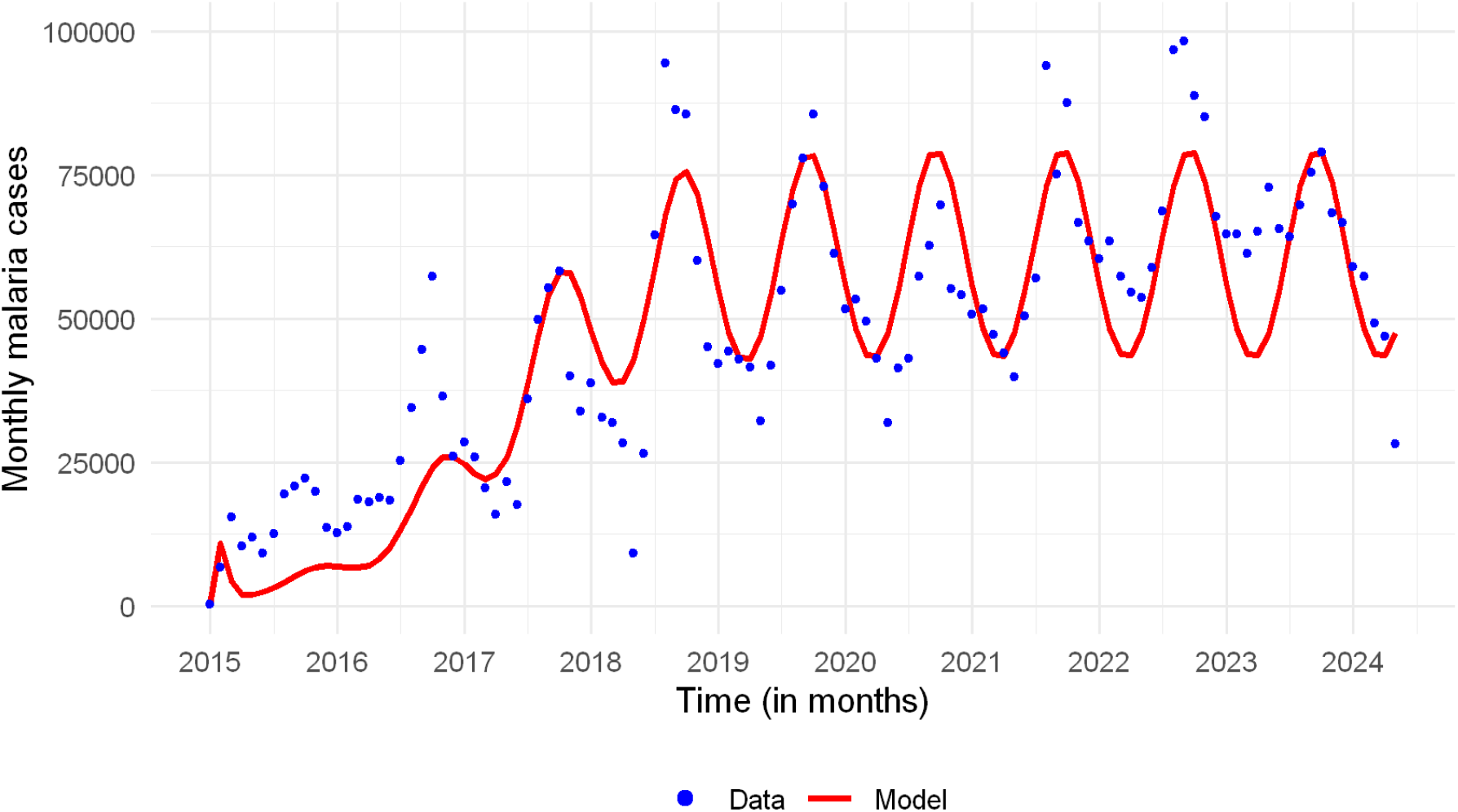
Plot showing monthly observed malaria cases in Kebbi State (dots) compared with the fitting output from model (10) (solid line)

Furthermore, the index of agreement (IOA) is used as a measure of the goodness-of-fit to quantify the association between model (10) and Kebbi State monthly malaria cases presented in 1b. The IOA is a widely used statistical metric for evaluating the performance of predictive models, particularly in environmental modelling contexts [40]. It quantifies how well model prediction matches observed data, considering both the magnitude and direction of deviations [40, 41]. The IOA originally proposed by [41] is mathematically expressed as

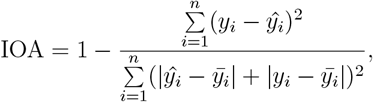

where *y*_*i*_ is the reported malaria cases, *ŷ*_*i*_ is the predicted values from the model (10), 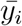 is the mean of reported malaria cases and *i* = 1, …, *n* (*n* is the number of observations). The value of IOA lies between 0 and 1, with 0 indicating no agreement and 1 indicating perfect agreement between the model and the data. For our model calibration, the IOA is 0.9308, which implies a strong degree of agreement between the model and the data.

### 2.5 Numerical simulations

We used the estimated parameters obtained through the model calibration carried out in Subsection 2.4, along with the fixed parameters in Table 3 and the initial conditions, to simulate malaria incidence in order to assess the effects of LLIN on malaria transmission in Kebbi State. We varied LLIN use (50% and 80%) relative to the baseline (38.2%), leading to a corresponding LLIN protective effectiveness *u*_1_ = 0.45, 0.72, and 0.34, respectively. This is relevant to providing evidence-based suggestions for the continuous rollout of LLINs and the need to gear up efforts to translate into greater LLIN use across communities in Kebbi State.

Specifically, numerical simulations of monthly malaria incidence are performed for retrospective analysis on the effects of LLIN on malaria transmission in Kebbi State. With the retrospective analysis, we quantified the magnitude of malaria cases that could have been averted under different scenarios in terms of variability in the use of LLIN. Furthermore, numerical simulations of future monthly malaria incidence are also performed considering the scenarios of 50% and 80% use of LLIN to assess the effects of LLIN on malaria transmission in Kebbi State.

## 3 Results

The results arising from numerical simulations of the malaria transmission dynamics and control model (10) are presented in this section.

In Table 4, the sensitivity indices of the effective reproductive number, ℛ_*e*_, with respect to the model parameters, using the baseline values of the parameters in Table 3, are presented.

**Table 4:** Sensitivity indexes of the effective reproduction number, ℛ_*e*_, to the parameters of malaria model (10)

| Parameter | Sensitivity index | Parameter | Sensitivity index |
| --- | --- | --- | --- |
| $\epsilon_0$ | +1 | $\theta_I$ | $-9.4300 \times 10^{-7}$ |
| $\beta_h$ | +0.50 | $\beta_m$ | +0.50 |
| $\mu_h$ | $-1.93542 \times 10^{-4}$ | $\sigma_m$ | +0.2083 |
| $\sigma_h$ | +0.00023 | $\Lambda_m$ | +0.50 |
| $\phi$ | +0.00002476 | $\Lambda_h$ | -0.50 |
| $\rho$ | +0.00003470 | $\mu_m$ | -1.2083 |
| $u_1$ | -0.5239 | $\delta$ | $-1.7258 \times 10^{-8}$ |
| $u_2$ | $-3.372 \times 10^{-5}$ | | |

It is observed from Table 4 that the model parameters are categorised into two according to the signs of their sensitivity indices – those that have positive sign of sensitivity index (*ϵ*_0_, *β*_*h*_, *σ*_*h*_, *ϕ, ρ, β*_*m*_, *σ*_*m*_, Λ_*m*_) and those that have negative sensitivity index (*µ*_*h*_, *δ, θ*_*I*_, Λ_*h*_, *µ*_*m*_, *u*_1_, *u*_2_). The most sensitive parameters in the first category are *ϵ*_0_, *β*_*h*_, *β*_*m*_ and Λ_*m*_, while the most sensitive parameters in the second category are *µ*_*m*_, *u*_1_ and Λ_*h*_. The implication of the parameters with positive sensitivity index is that the value of the effective reproduction number, ℛ_*e*_, will increase (or decrease) when the value of any of the parameters in this category is increased (or decreased). For example, the normalised sensitivity index of ℛ_*e*_ with respect to *ϵ*_0_, 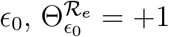 suggests that the value of the effective reproduction number, ℛ_*e*_, will increase by 10% when the value of the per capita biting rate of mosquitoes, *ϵ*_0_, increases by 10%. Similarly, the negative sign of the effective reproduction number, ℛ_*e*_ to the model parameters suggests that an increase in the value of any of the parameters in this category will lead to a decrease in ℛ_*e*_ value. For example, 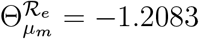 suggests that 10% increase (or decrease) in the baseline value of the natural mortality rate of mosquitoes, *µ*_*m*_, corresponds to 12.083% decrease in the value of ℛ_*e*_. We further focus on the most sensitive model parameters - per capita mosquito biting rate (*ϵ*_0_), protective effectiveness of LLINs (*u*_1_), transmission probabilities in human and mosquito populations (*β*_*h*_ and *β*_*m*_, respectively) and natural mortality rate of mosquitoes (*µ*_*m*_) - and numerically illustrate how they influence the dynamics of malaria transmission. See the results in Figures 4 and 5.

**Figure 4.**
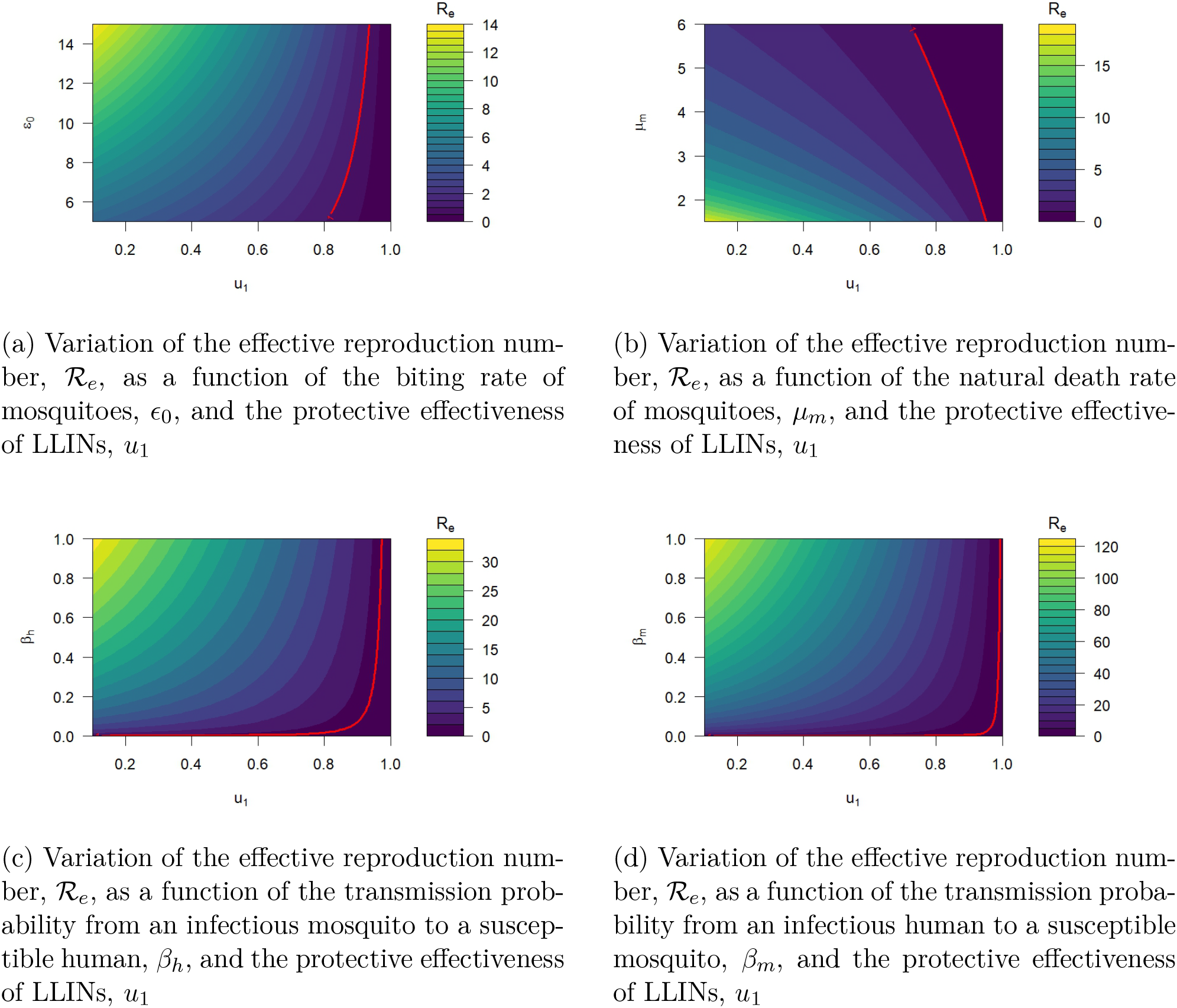
Heat map showing the variations of the effective reproduction number, ℛ_*e*_ with respect to different combinations of key sensitive parameters with the protective effectiveness of LLINs (*u*_1_). The colour gradient represents the magnitude of ℛ_*e*_, while the red contour line denotes the epidemic threshold (ℛ_*e*_ = 1), separating regions of disease persistence (ℛ_*e*_ > 1) from elimination (ℛ_*e*_ < 1)

**Figure 5.**
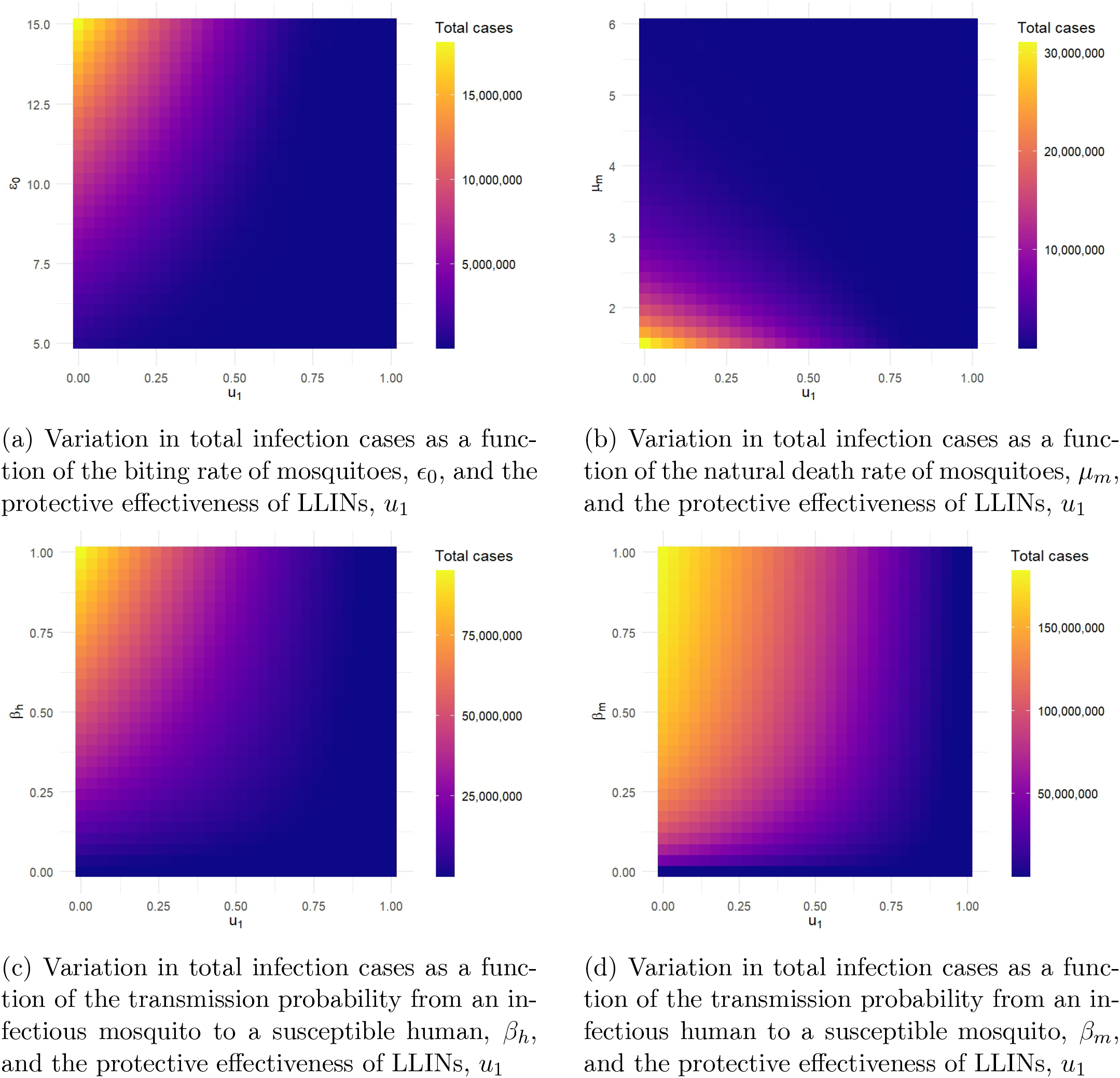
Heat map showing the variations in total infection cases with respect to different combinations of key sensitive parameters with the protective effectiveness of LLINs (*u*_1_). The color gradient represents total cumulative cases

Figure 6 illustrates the predicted monthly incidence of malaria in Kebbi State for a period of nine years and five months (January 2015 to May 2024) with three different levels of of LLIN usage: 38.2%, 50% and 80%, describing what the malaria transmission dynamics could have been under these different usage of LLINs. The baseline usage of LLIN is 38.2% obtained from the National Malaria Indicator Survey (NMIS). Figure 7 describes the simulated result of the predicted monthly malaria cases under different usage of LLINs (38.2%, 50%, and 80%), which spans 5 years. Table 5 highlights the cases of malaria averted in relation to the baseline usage of LLINs using the other usage level as scenarios.

**Table 5:** Results of the estimated number of averted malaria cases under different usage levels of LLINs relative to the baseline usage.

| Scenarios | Usage level of LLINs | Relative cases averted | Percentage case averted(%) |
| --- | --- | --- | --- |
| A | 50% | 1,980,288 | 37.89 |
| B | 80% | 5,121,122 | 97.98 |

**Figure 6.**
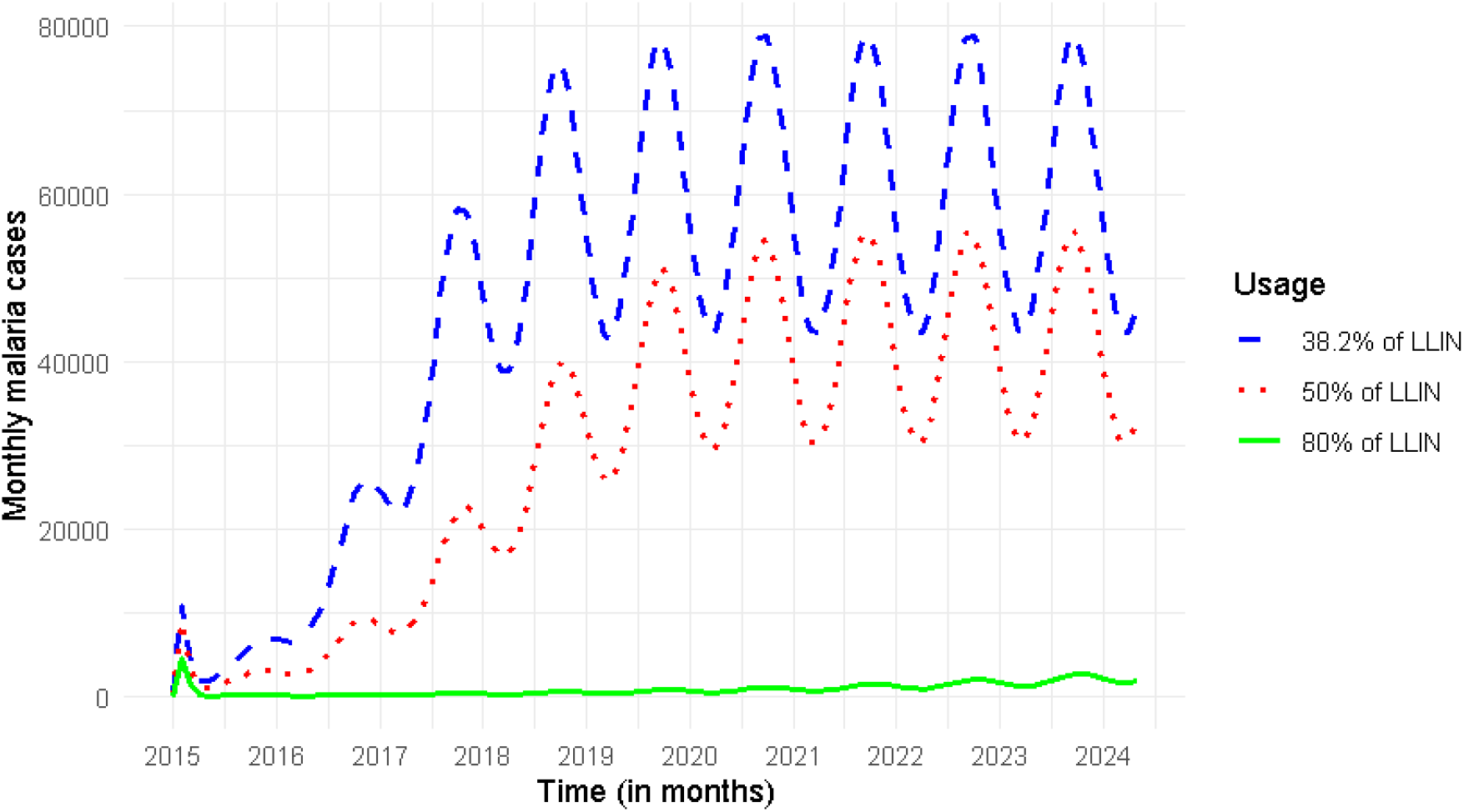
Simulated malaria case trajectories in Kebbi State, Nigeria, under different scenarios of LLINs usage. The scenarios illustrate how variations in LLINs usage influence malaria transmission dynamics

**Figure 7.**
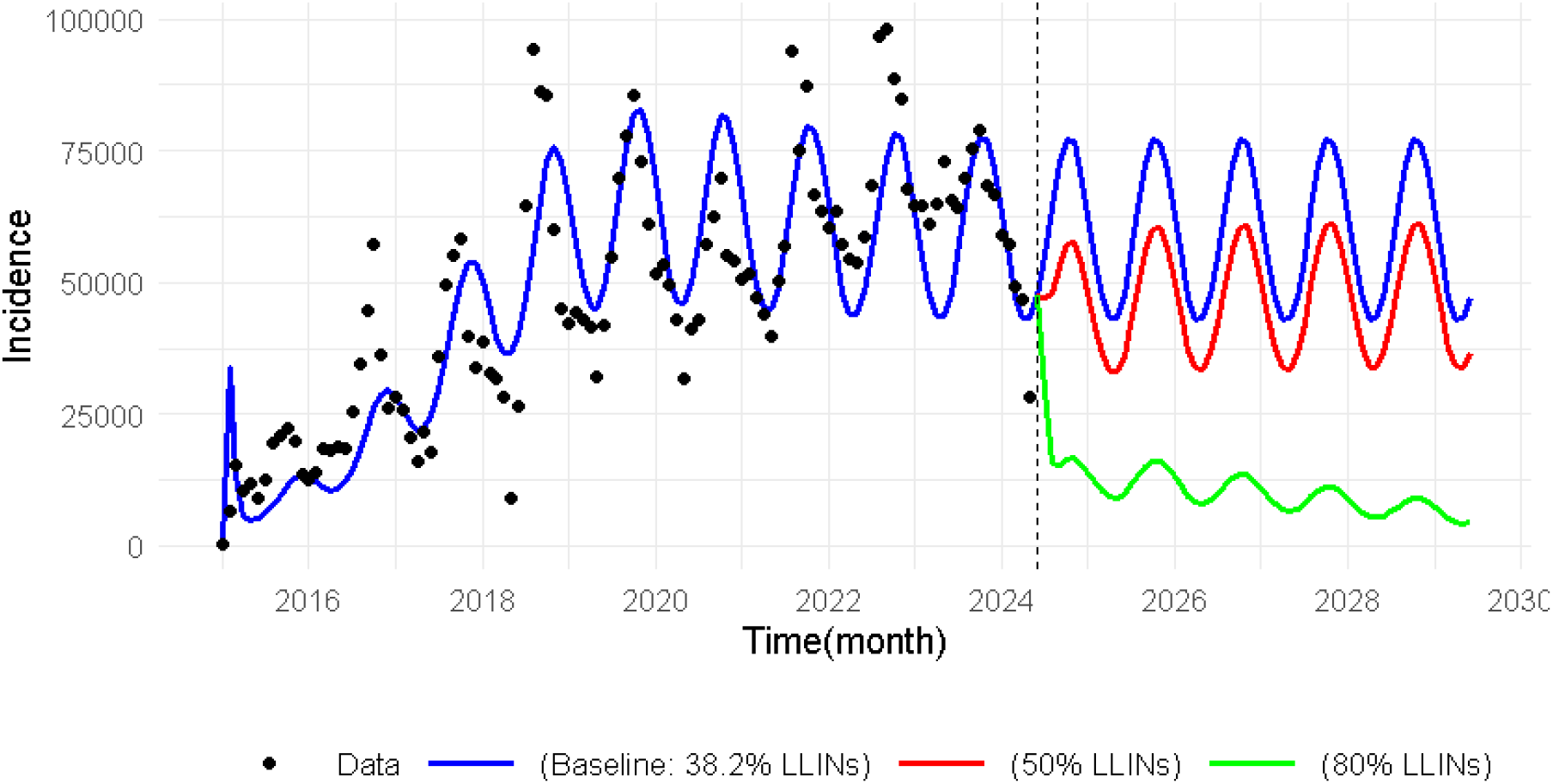
Model based predictions of malaria cases in Kebbi State, Nigeria, from 2024 to 2029 under different levels of LLINs usage, highlighting the influence of LLIN usage on future malaria transmission dynamics

## 4 Discussion

In this section, a detailed discussion of the results is considered. The results of the local sensitivity analysis summarised in Table 4 offer insightful information about how the protective effectiveness of LLINs, *u*_1_, influences the transmission dynamics and control of malaria in the interacting populations of human and mosquito. In particular, a 10% increase in the protective effectiveness of LLINs will lead to approximately a 5% reduction in the effective reproduction number. Therefore, intensifying the efforts on LLINs usage may be helpful towards malaria elimination in the population.

The plots in Figure 4 demonstrate how variations in the most sensitive parameters - per capita biting rate of mosquitoes (*ϵ*_0_), natural mortality rate of mosquitoes (*µ*_*m*_), transmission probability from an infectious mosquito to a susceptible human (*β*_*h*_), transmission probability from an infectious human to a susceptible mosquito (*β*_*m*_) - influence the effective reproduction number (ℛ_*e*_), thereby impacting the speed of disease spread. However, higher levels of protective effectiveness of LLINs (*u*_1_) significantly suppress ℛ_*e*_, shifting the system towards the disease-free region. The steep transition around the red threshold line indicates that a modest upscale in LLIN effectiveness near the critical level can effectively drive ℛ_*e*_ below one, leading to the disease elimination. This highlights that maintaining strong protective effectiveness of LLINs can offset high transmission potential and prevent resurgence even when the probabilities of transmission and mosquito biting rates are moderately high and the natural mortality rate of mosquitoes is moderately low.

Moreover, the incidence heat map in Figure 5 exhibits a similar trend to the reproduction number pattern. On the one hand, total cases rise sharply with increasing mosquito biting rate (*ϵ*_0_) and drop significantly as protective effectiveness of LLINs (*u*_1_) strengthens as shown in Figure 5a. Similar observations are made for increases in the probabilities of transmission in humans and mosquitoes (*β*_*h*_ and *β*_*m*_, respectively), where the total infection cases even reach their peaks. On the other hand, total cases drop sharply with increasing natural mortality rate of mosquitoes and increasing protective effectiveness of LLINs, *u*_1_ as demonstrated in Figures 5b, 5c and 5d, respectively. The steep gradient from yellow to blue indicates that even moderate control levels can substantially reduce disease burden. This demonstrates that moderate improvements in LLIN effectiveness can produce large reductions in total cases, emphasising the importance of sustained LLIN usage in reducing malaria incidence.

Figure 6 provides information on how variation in LLIN usage between January 2015 and May 2024 could have reduced malaria incidence cases in Kebbi State. By comparing these scenarios at 50%, and 80% levels of usage, we can better understand the role of LLIN in reducing malaria transmission in Kebbi State. The blue line represents the baseline scenario with 38.2% LLIN usage, reflecting the usage of LLIN in Kebbi State as of 2015, meanwhile, NMEP proposed increasing their coverage level (same as usage in the context of this paper) to 80% by 2025 [9]. These figures formed the choice of usage level adopted for simulation for this study. The red line, which corresponds to 50% LLIN usage, shows a decrease in malaria incidence compared to the baseline. In this scenario, the incidence of malaria cases remains high throughout the period, indicating that with low usage, a large percentage of the population remains unprotected, allowing continuous mosquito-human transmission due to the limited impact of LLIN usage. This implies that, by increasing usage by only 11.8%, the number of malaria cases reduced with an estimated 1,980,288 cases averted, equating to a 37.89% reduction relative to the 38.2% usage level.

This outcome shows the benefit of moderate improvements in LLIN usage, illustrating scaling LLIN usage to half of the population could achieve 37.89% malaria cases averted in Kebbi State (see Table 5). This level of reduction emphasises improvement achievable through intermediate LLIN usage goals, especially in settings where full LLIN adoption may be challenging to implement immediately. The green line, which represents the plot of the highest usage level at 80%, demonstrates the most dramatic effect of LLIN on malaria incidence. At this usage level, malaria cases decrease rapidly over the period, nearing elimination in the latter years. Compared to the baseline, 80% LLIN usage is expected to avert 5,121,122 malaria cases, amounting to a 97.98% reduction in malaria cases, as seen in Table 5. This finding shows the potential effect of high usage in effectively reducing malaria transmission, suggesting that malaria could be brought under control within a decade if a sustained 80% usage level is achieved and maintained. Such significant reduction provides compelling evidence that high LLIN usage could serve as a basic strategy for malaria elimination in Kebbi State and similar endemic regions.

In addition, the result in Figure 7 illustrates the prediction of malaria incidence in Kebbi State from June 2024 to December 2029, with respect to different scenarios under consideration. Before this point, malaria incidence followed a historical trajectory, represented by the blue plot in Figure 7, which reflects a “business as usual” scenario with a baseline LLIN usage of 38.2%. After May 2024, we implemented two additional LLIN usage levels: 50% and 80%. By integrating these different usage levels into our model, we simulated their effects on malaria incidence and visualized the results alongside the historical data, represented by black dots. This allowed us to effectively compare the actual observed incidence with our model predictions. To quantify the impact of these increased LLIN usage levels, we calculated the total predicted incidence for each scenario. The findings revealed that with a baseline usage of 38.2%, the predicted incidence was approximately 3,681,628 cases as displayed in Table 6. In contrast, with a 50% usage level, the incidence decreased to about 2,643,205 cases, while an 80% usage level led to a further reduction to approximately 775,551 cases. From the analyse, an increase to 50% usage is predicted to avert approximately 1,038,423 malaria cases, which corresponds to a 28.21% reduction in predicted incidence. Moreover, scaling up usage to 80% could avert about 2,906,077 cases, signifying an impressive reduction of 78.93% in incidence (see Table 6).

**Table 6:** Results of the estimated number of averted malaria cases for 5 years prediction under different usage levels of LLINs relative to the baseline usage.

| Scenarios | usage level of LLINs | Total cases | Relative cases averted | Percentage case averted(%) |
| --- | --- | --- | --- | --- |
| Baseline | 38.2% | 3,681,628 | - | - |
| A | 50% | 2,643,205 | 1,038,423 | 28.21 |
| B | 80% | 775,551 | 2,906,077 | 78.93 |

These findings align with the results reported by some previous studies [42, 43, 44, 45], supporting the effectiveness of LLINs in reducing malaria incidence. For instance, the study in [44] revealed that the use of LLIN led to a 50% reduction in malaria incidence in sub-Saharan Africa. Consequently, these results align with global malaria control strategies endorsed by WHO [7], which recommended scaling up the distribution of LLIN and other preventive measures in high-burden settings [46]. This paper supports the United Nations’ Sustainable Development Goal 3 (SDG 3), which seeks to ensure healthy lives and promote well-being for all. In particular, we focus on target 3.3, which aims to eliminate malaria and other diseases by 2030. In line with SDG 3, this study contributes to current malaria control efforts and will help to improve the distribution of LLINs in Kebbi State.

Our findings reveal that while LLIN usage is currently at 38.2%, malaria cases remain high, a clear indication that many individuals are either not receiving nets or, more critically, not using them consistently. To effectively reduce malaria transmission in Kebbi State, public health policymakers must urgently refocus efforts on increasing the actual usage of LLINs, not merely their distribution. The implications of our results for public health policy in Kebbi State highlight that increasing LLIN usage not only protects more people but also helps kill mosquitoes, reducing their lifespan and ability to transmit the disease. This explains the trends we see in our simulation results and point to clear actions: reduce mosquito bites through prevention, increase mosquito death rates through vector control, and expand LLIN usage as widely as possible. The result also offers practical insights for planning and resource allocation. Reaching 50% LLIN usage is a realistic short-term goal that can still make a big difference, especially in areas with limited resources. In the long term, aiming for 80% usage should be a top priority. Even modest improvements in usage can yield dramatic health benefits. These predictions reinforce the importance of LLINs as a frontline tool in Nigeria’s malaria elimination strategy.

Although LLIN distribution campaigns have been implemented in the past, there is a pressing need to assess whether these nets are being used effectively. Mass replacement of expired or damaged LLINs should be prioritized, especially in communities where usage has declined due to wear and tear or misconceptions. Distribution efforts must be paired with planned follow-up mechanisms to ensure that nets are not only delivered but also installed and used correctly. Community health workers should be trained and deployed to conduct household visits, demonstrate proper net hanging techniques, and monitor usage patterns over time.

Equally important is addressing the behavioural and cultural barriers that hinder LLIN usage. Many individuals avoid using nets due to discomfort, heat, or unfounded fears [6]. To overcome these challenges, literacy and orientation campaigns must be intensified. These should be delivered in local languages and tailored to community norms, using trusted voices such as religious leaders, traditional rulers, and local influencers. Radio programs, street theatre, and mobile messaging can be powerful tools to normalise LLIN usage and dispel myths. Where such efforts are already in place, they must be sustained and scaled; where absent, they should be urgently introduced.

Despite the progress that could be made due to the use of LLINs as the findings of this study have revealed, potential barriers such as inconsistent use of nets, mosquitoes developing resistance to insecticides, and difficulties in getting nets to remote communities can limit the success of LLIN in preventing malaria incidence in Kebbi State. Addressing these issues is essential to fully achieve the benefits shown in our results.

Finally, policymakers must invest in monitoring and evaluation systems that track not just LLIN usage, but actual usage rates and their impact on malaria incidence. Real-time data dashboards, periodic household surveys, and community feedback loops are essential for refining strategies and ensuring accountability. Continued political commitment, adequate funding, and community ownership will be vital to sustaining progress. The evidence is clear: increasing LLIN usage, not just usage, is the key to unlocking major health gains and moving Kebbi State closer to malaria elimination. With the right investments and strategies, Kebbi State can make major progress toward eliminating malaria.

## 5 Conclusion

Long-lasting insecticidal nets (LLINs) are a major control intervention tools for the fight against malaria in Nigeria. In this study, a deterministic compartmental model of malaria that incorporates the effects of LLINs was fitted to the monthly routine data for Kebbi State, Nigeria from January 2015 to May 2024. To establish the epidemiological feasibility of the malaria model, we performed some qualitative analysis such as positivity and boundedness of solutions. The effective reproduction number was computed, and the stability of equilibria under certain conditions was analysed. Model fitting and parameter estimation were performed using the least squares method. Some of the parameters were obtained from existing literature while others were derived from demographic data. The effective reproduction number ℛ_*e*_ was estimated to be 7.39.

Local sensitivity analysis was performed to determine the most influential parameters driving malaria transmission in Kebbi State. The study showed that in comparison to the baseline usage, 50% LLIN usage will lead to reduction of malaria cases by 37.89% while 80% LLIN usage will reduce malaria cases by 97.98%. We also carried out scenario analysis to make a 5-year prediction of malaria incidence with varying levels of LLINs usage: 38.2% (baseline usage), 50% and 80%. Compared to baseline usage of 38.2%, the result shows that scaling up the usage of LLINs to 50% in Kebbi State will reduce the incidence of malaria by 28.21%, while scaling up to the level of usage 80% will lead to a significant reduction of 78.93%.

However, the study is not without some limitations. Firstly, we have assumed a constant LLINs protective effectiveness rate and have not accounted for the decay rate of LLINs. This assumption in our model could lead to over-estimation of LLIN protective effectiveness in reducing malaria cases in the model predictions. Thus, future studies intend to incorporate decay functions to capture the time dependent loss of LLIN efficacy. Secondly, to reduce model complexity, the study did not explicitly account for the additional killing effect of LLINs. It is therefore recommended that, to bring down malaria incidence in Kebbi State to its bearest minimum, health policymakers in Nigeria should prioritise scaling up LLINs usage in the State by expanding LLIN distribution in the State and creating awareness on its proper usage.

## Data Availability

All data produced in the present work are contained in the manuscript

https://nmdrnigeria.ng/dhis-web-commons/security/login.action

## Appendix A

**Qualitative analysis of the malaria model**

In this subsection, we take up the qualitative analysis of the autonomous version of the proposed malaria model by setting the seasonal-forcing mosquito biting rate *ϵ*(*t*) = *ϵ*_0_.

### Appendix A.1 Boundedness of solutions

#### Theorem A.1.

*Let N*_*h*_(*t*) *and N*_*m*_(*t*) *represent the total human and mosquito populations in system* (10), *respectively, which satisfies the differential equations:*

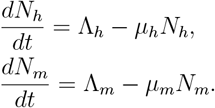

*Then, the total populations N*_*h*_(*t*) *andN*_*m*_(*t*) *are uniformly bounded if:*

1. 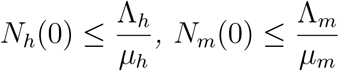, *than* 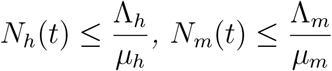 *for all time t >* 0;
2. 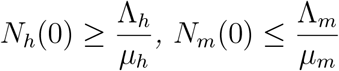 *than N*_*h*_(*t*) *andN*_*m*_(*t*) *asymptotically approach* 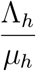 *and* 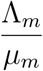, *respectively, as t* → ∞.

#### Proof.

The rate of change of the total human population is obtained by adding the right hand side of the first five equations in system (10):

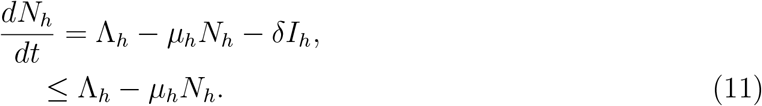

Integrating the inequality in (11) yields,

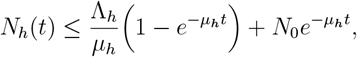

so that, as *t* → ∞, the human population is bounded by

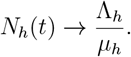

Similarly, the rate of change of the total mosquito population is obtained by adding the right-hand side of the last three equations in system (10) as

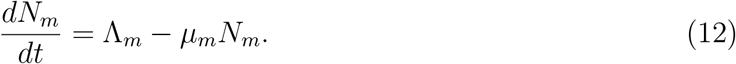

It follows from integrating the result in (12) that

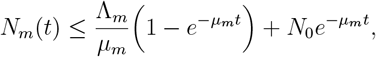

such that, as *t* → ∞, the mosquito population is bounded by

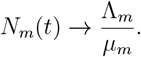

Since *µ*_*h*_ > 0, if 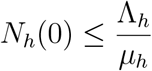, then 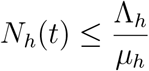 for all *t >* 0. This proves that *N*_*h*_ (*t*) is bounded above by 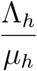. Also, since *µ*_*m*_ > 0, if 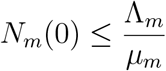, then 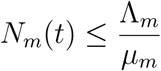 for all *t >* 0. This proves that *N*_*m*_ (*t*) is bounded above by 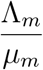. Therefore, both human, *N*_*h*_ (*t*), and mosquito, *N*_*m*_(*t*), populations remain within biologically meaningful bounds. Hence, the proof.

### Appendix A.2 Positivity of solutions

To ensure that the solutions remain biologically meaningful, we now prove that all state variables remain non-negative for all time *t* > 0.

#### Theorem A.2.

*Suppose the initial values for the state variables are given by*

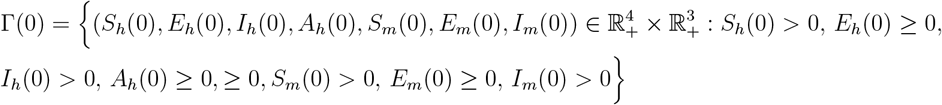

*Then, the solutions of the system* (10) *satisfy*

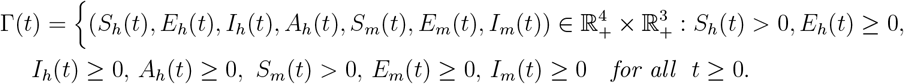

#### Proof.

Considering the system in 10, we have

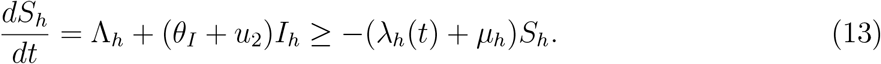

Integrating 13 gives

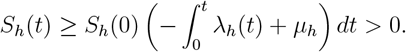

Similarly in system 10, it can be shown that, *E*_*h*_(*t*) > 0, *I*_*h*_(*t*) > 0, *A*_*h*_(*t*) > 0, *S*_*m*_(*t*) > 0, *E*_*m*_(*t*) > 0, and *I*_*m*_(*t*) > 0 for all *t* > 0.

We have shown that the solutions of the model are positive for all time *t* > 0 provided that the initial values of the state variables are non-negative. These results establish the biological feasibility of the model.

#### Appendix A.2.1 Disease free equilibrium point

The disease-free equilibrium point (DFE) occurs when the system reaches a steady state with no malaria infections (i.e., no infected individuals exist in the population). Specifically, the DFE of system (10) occurs when

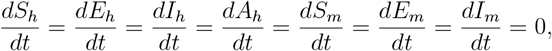

with *E*_*h*_ = *I*_*h*_ = *A*_*h*_ = *E*_*m*_ = *I*_*m*_ = 0.

Now, from the first differential equation of model 10 at equilibrium, we have

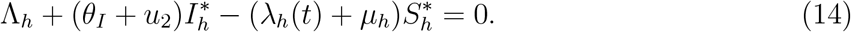

Solving for 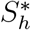 in 14 gives

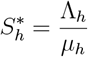

Similarly, it is easy to obtain the equilibrium solution of the fifth equation of the malaria model 10 as

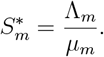

Therefore, DEF of the system 10 is obtained as

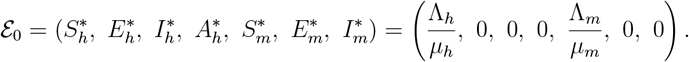

### Appendix A.3 Effective reproduction number

The effective reproduction number, denoted as ℛ_*e*_, quantifies the potential for infection transmission in a population in the presence of interventions. Using the next-generation method, the van den Driessche and Watmough approach outlined in [47] is employed to compute the next-generation matrix related to model (10). The model is then written as 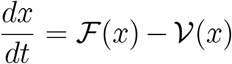 where ℱ represents the rate of appearance of new infections, and 𝒱 describes the transfer of individuals in and out of the infected compartments.

Let the dynamics of the infected compartments (*E*_*h*_, *I*_*h*_, *A*_*h*_, *E*_*m*_, *I*_*m*_) of model (10) be represented as

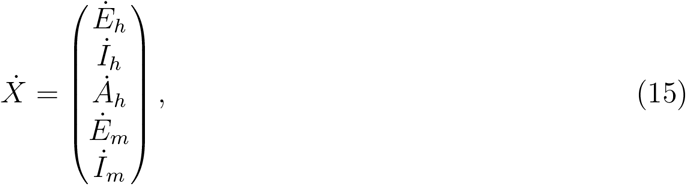

and the linearized system at the DFE takes the form

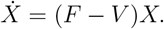

Then, the rate of appearance of new infections in the population, *f*_*i*_, is given by

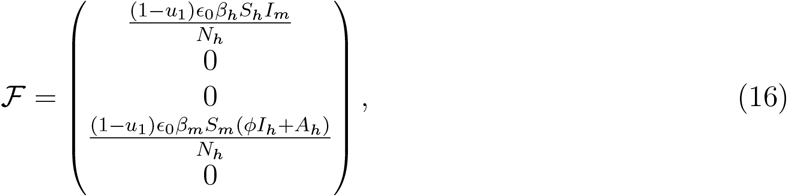

while the rate of transfer of individuals in and out of the infected compartments, *v*_*i*_, is given by

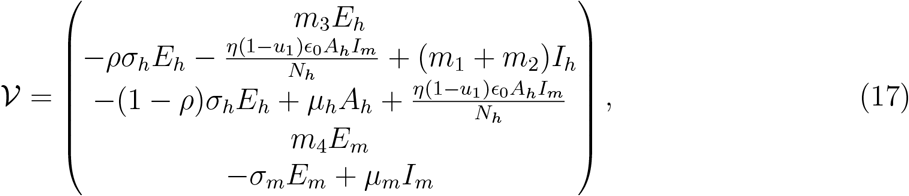

where *m*_1_ = *θ*_*I*_ + *u*_2_, *m*_2_ = *δ* + *µ*_*h*_, *m*_3_ = *σ*_*h*_ + *µ*_*h*_ and *m*_4_ = *σ*_*m*_ + *µ*_*m*_. Hence, finding the Jacobian matrix of F in (16) and the Jacobian matrix of V in (17) at the DFE gives

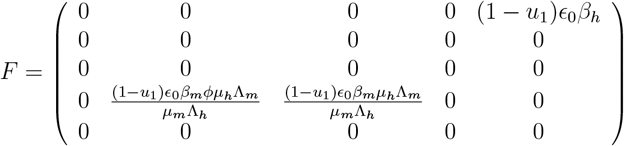

and

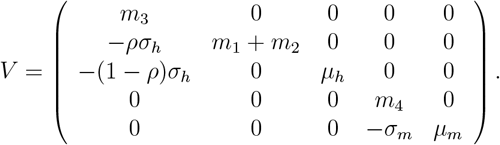

It follows that

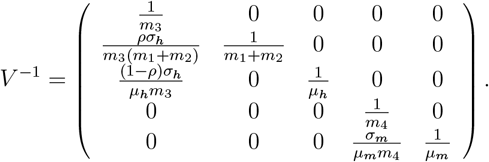

The next generation matrix *FV* ^−1^ is obtained as

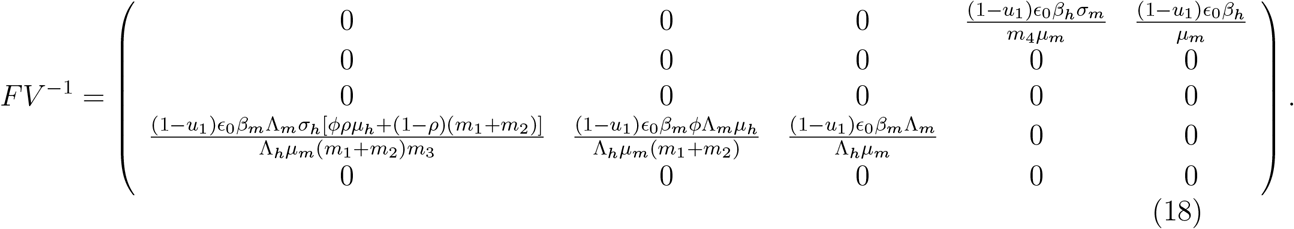

Therefore, the effective reproduction number of the malaria model (10) is the largest eigenvalue (also called the *spectral radius*) of *FV* ^−1^ in (18), and is obtained as

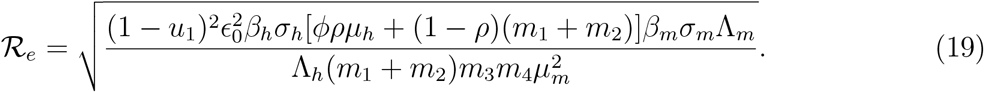

If ℛ_*e*_ > 1, the disease spread in the population which might lead to an outbreak or an epidemic. Also, if ℛ_*e*_ < 1, the infection will die out over time as each infected individual produces on average fewer than one new infection.

### Appendix A.4 Local stability of disease free equilibrium

The existence of a disease-free equilibrium (DFE) has been established for the malaria model. To determine the long-term behavior of this equilibrium, we perform a stability analysis to assess whether the DFE is locally asymptotically stable or unstable.

#### Theorem A.3.

*The malaria disease-free equilibrium state of the model is locally asymptotically stable when* ℛ_*e*_ < 1 *and unstable when* ℛ_*e*_ > 1.

#### Proof.

Jacobian matrix of model (10) at DFE is given by

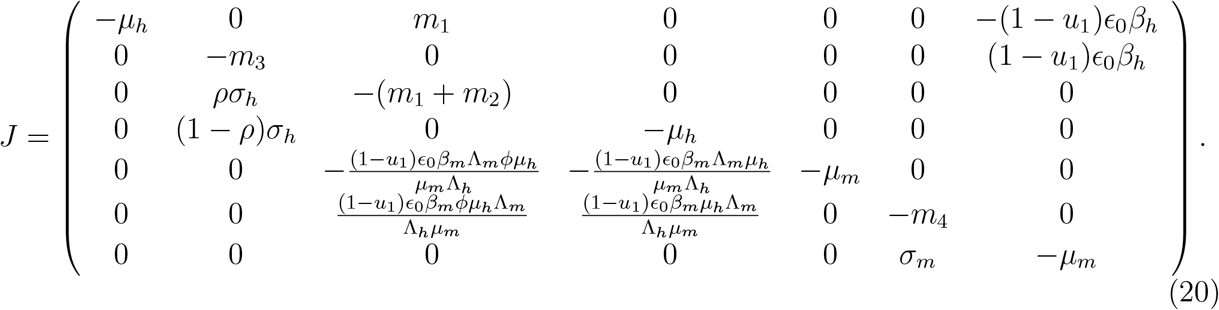

The Jacobian matrix (20) provides seven eigenvalues, which include − *µ*_*h*_ and − *µ*_*m*_. The remaining eigenvalues are the roots of the characteristic equation

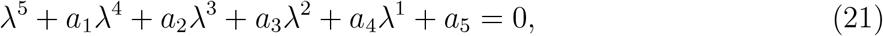

where

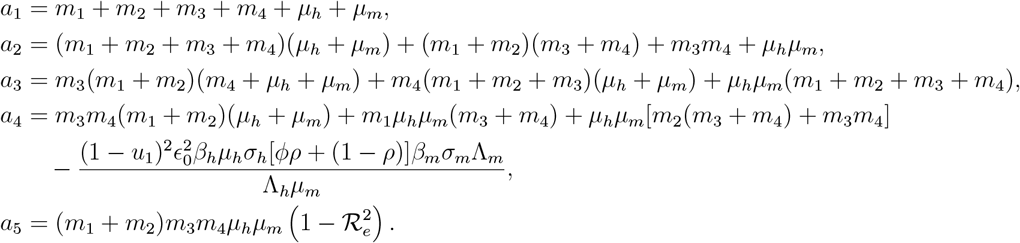

By the Routh-Hurwitz criteria in [48], the polynomial in (21) has negative real roots provided that 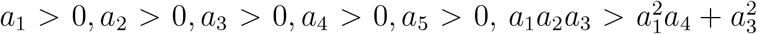 and 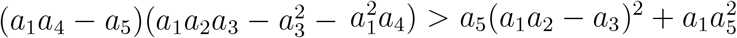 along with the condition ℛ_*e*_ < 1. It implies that, with negative real eigenvalues, the disease-free equilibrium state is locally asymptotically stable.

### Appendix A.5 Endemic equilibrium point of model (10)

At the endemic equilibrium, the infection persists in the population, so *E*_*h*_, *A*_*h*_, *I*_*h*_, *E*_*m*_, *I*_*m*_ ≠ 0 in the malaria model (10). Let the endemic equilibrium of the model be given by

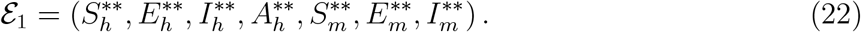

Further, let the forces of infection in human and mosquito at steady state be defined as

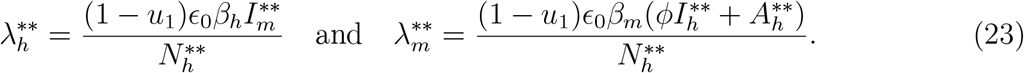

Thus, solving system (10) at steady state in terms of the forces of infection in (23) gives the components of ℰ_1_ in (22) as

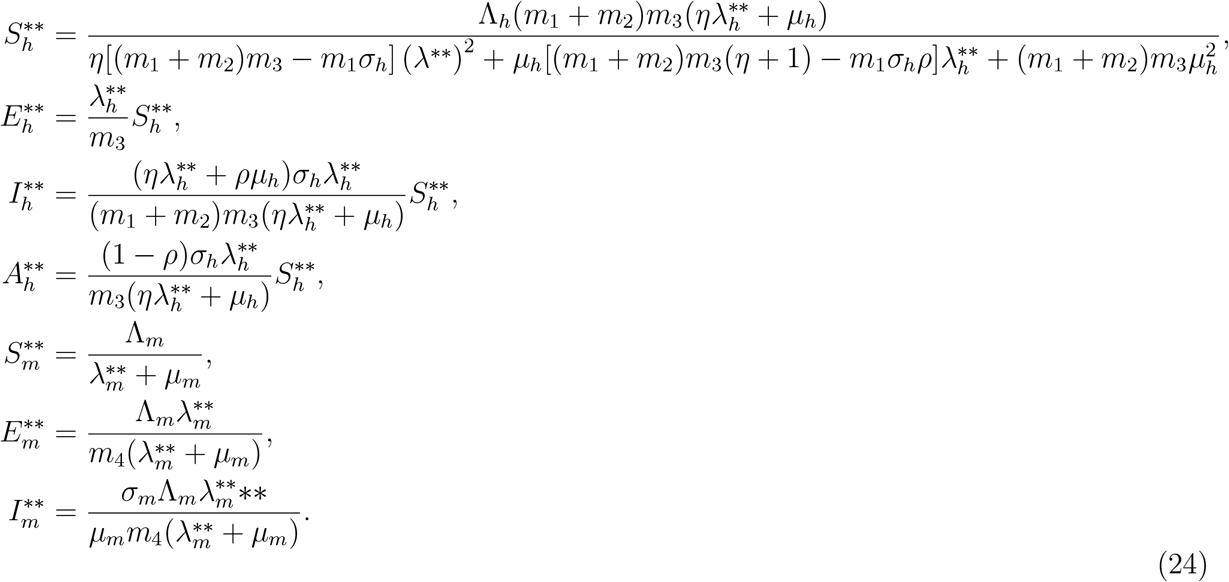

Substituting the appropriate results in (24) into the forces of infection in (23) and simplifying give rise to the following polynomial equation:

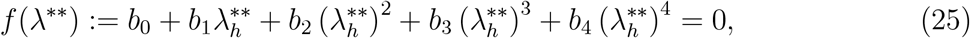

where

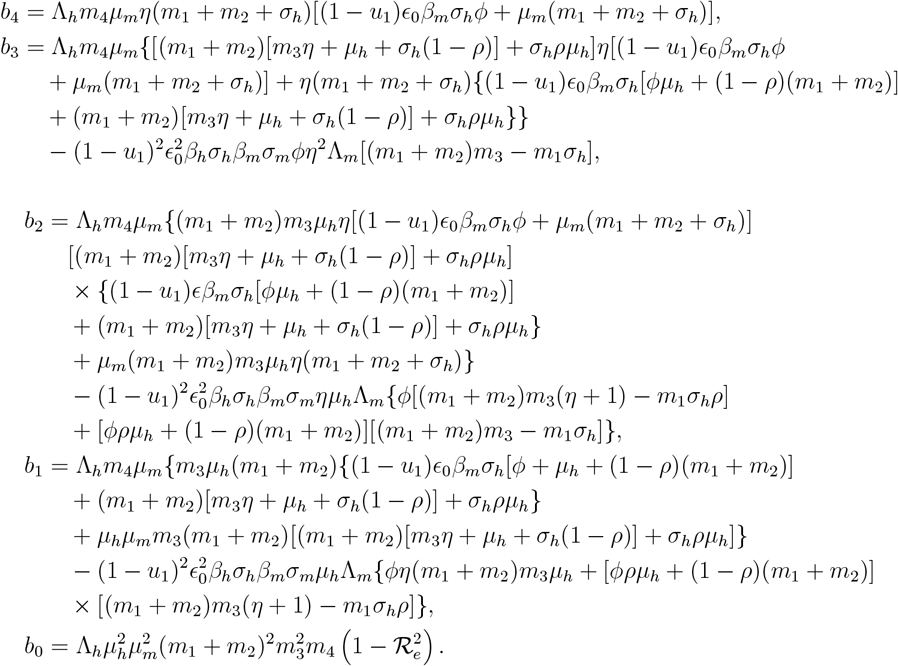

Solving for 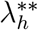 in (25) consequently leads to obtaining the positive endemic equilibrium ℰ_1_ of the malaria model (10). Since the model parameters are non-negative, *b*_4_ > 0 always holds, whereas *b*_0_ > 0 (*b*_0_ < 0) whenever ℜ_*e*_ < 1 (ℜ_*e*_ > 1). Thus, the number of possible positive real roots of polynomial (25) is determined by the signs of the coefficients *b*_1_, *b*_2_ and *b*_3_. Solving (25) for the positive values of 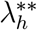 allows us to explicitly determine the positive endemic equilibria of the malaria model (10). By Descartes’ rule of signs (see Theorem 4.10 in [49]), the summary of the various possibilities for the positive roots of the polynomial whenever ℜ_*e*_ < 1 and ℜ_*e*_ > 1 are displayed in Table 7.

**Table 7:** Different cases and their corresponding total number of possible real roots of polynomial (25)

| Case | Coefficient of polynomial (25) | | | | | $\mathcal{R}_e$ | Number of sign changes | Number of possible positive real roots |
| --- | --- | --- | --- | --- | --- | --- | --- | --- |
| | $b_0$ | $b_1$ | $b_2$ | $b_3$ | $b_4$ | | | |
| I | + | + | + | + | + | $< 1$ | 0 | 0 |
| II | - | - | + | + | + | $> 1$ | 1 | 1 |
| III | + | + | - | + | + | $< 1$ | 2 | 0, 2 |
| IV | - | + | + | - | + | $> 1$ | 3 | 1, 3 |
| V | + | - | + | - | + | $< 1$ | 4 | 0, 2, 4 |

It should be mentioned that all the other possible sign changes such that Cases II, III and IV hold are omitted in Table 7. Consequently, the results in the table give birth to the following Theorem:

#### Theorem A.4.

*The model of malaria transmission dynamics and control* (10) *admits:*

i. *Only one (unique) endemic equilibrium when case II in Table 7 is applicable and* ℜ_*e*_ > 1;
ii. *More than one endemic equilibrium when case IV in Table 7 is applicable and* ℜ_*e*_ > 1;
iii. *More than one endemic equilibrium if cases III and V in Table 7 are applicable such that* ℜ_*e*_ < 1;
iv. *No endemic equilibrium otherwise, whenever case I in Table 7 holds and* ℜ_*e*_ < 1.

### Appendix A.6 Sensitivity analysis

In this part of the paper, the focus is on examining how the model parameters influence the transmission dynamics and control of *Plasmodium falciparum* malaria in the interacting human and mosquito populations. This paper considers local sensitivity analysis. In that case, the sensitivity indices of the effective reproduction number, ℜ_*e*_, with respect to the model parameters are obtained using the normalized forward-sensitivity index defined by

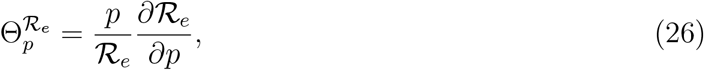

where *p* is any parameter contained in the expression for ℜ_*e*_ in (19). Thus, we are led to the following analytical sensitivity indices:

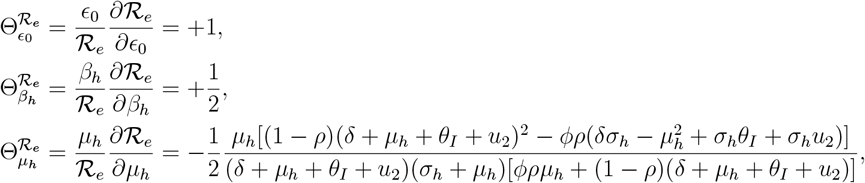

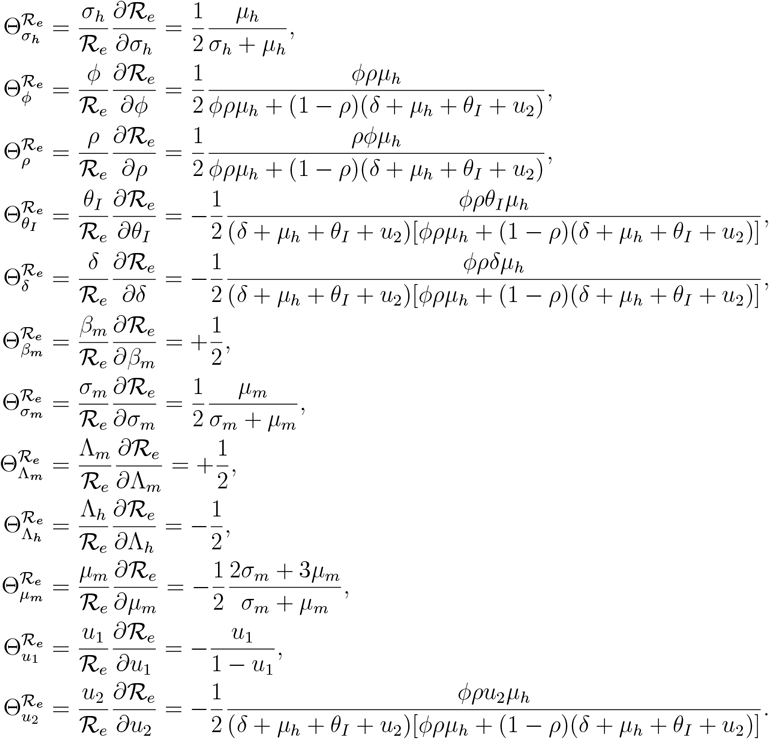

All the analytical indices derived, as presented above, are quantified in Section 3.

## CRediT authorship contribution statement

**Emmanuel A. Bakare**: Review and editing, Validation, Methodology, Investigation, Formal analysis, Conceptualization, Data and fund acquisition, Supervision, Software. **Idowu I. Olasupo**: Writing–original draft, Review and editing, Validation, Methodology, Investigation, Formal analysis, Software. **Micheal Imoudu**: Writing–original draft, Review and editing, Validation, Methodology, Investigation, Formal analysis. **Afeez Abidemi**: Writing–original draft, Review and editing, Validation, Methodology, Investigation, Formal analysis. **Deborah O. Daniel**: Writing–original draft, Review and editing, Validation, Methodology, Investigation, Formal analysis. **Samuel A. Osikoya**: Writing–original draft, Review and editing, Validation, Methodology, Investigation, Formal analysis. **Oluwaseun A. Mogbojuri**: Writing–original draft, Review and editing, Validation, Methodology, Investigation, Formal analysis. **Aaron O. Nwana**: Writing–original draft, Review and editing, Validation, Methodology, Investigation, Formal analysis. **Dolapo O. Oniyelu**: Writing–original draft, Review and editing, Validation, Methodology, Investigation, Formal analysis. **Ronke D. Olorunfemi**: Writing–original draft, Review and editing, Validation, Methodology, Investigation, Formal analysis. **Samson O. Olagbami**: Writing–original draft, Review and editing, Validation, Methodology, Investigation, Formal analysis. **Dorcas O. Agboola**: Writing–original draft, Review and editing, Validation, Methodology, Investigation, Formal analysis. **Steven I. Ikediashi**: Review and editing, Investigation, Formal analysis. **Ayomide Adedeji**: Review and editing, Investigation, Formal analysis. **Oluwaponmile Ikuseka**: Review and editing, Investigation, Formal analysis. **Micheal Fadairo**: Review and editing, Investigation, Formal analysis. **Odunayo Odewale**: Review and editing, Investigation, Formal analysis. **Goodness Onwuka**: Review and editing, Investigation, Formal analysis. **Aurel Hansinon**: Review and editing, Investigation, Formal analysis. **Dolapo A. Bakare**: Review and editing, Investigation, Formal analysis. **Lisa J. White**: Review and editing, Investigation, Formal analysis, Validation. **Nakul Chitnis**: Review and editing, Investigation, Formal analysis, Validation. **Olusola Oresanya**: Review and editing, Investigation, Formal analysis, Validation. **Chukwu Okoronkwo**: Review and editing, Investigation, Formal analysis, Data acquisition. **Eze Nelson**: Review and editing, Investigation, Formal analysis, Data acquisition. **Segun Kosoko**: Review and editing, Formal analysis. **Gabriel I. Ogban**: Review and editing, Formal analysis.

## Acknowledgments

The authors are grateful to the anonymous reviewers for their constructive comments, which have improved the quality of the original manuscript, and to NMEP and Malaria Consortium for their resourceful support.

## Declaration of competing interest

The authors declare that they have no conflict of interest.

## Code and data availability

The R codes used to generate the results presented in this manuscript are available from the corresponding author upon a reasonable request. Interested readers can reach out to the National Malaria Elimination Programme (NMEP), Nigeria, for the data used for this study.

## Funding statement

This work was supported by a grant from the Gates Foundation (INV-047051). The funders had no role or influence on the design and interpretation of the data collected, as well as in writing the manuscript.

## Notes

### Competing Interest Statement

There is no competing interests

### Funding Statement

This study was funded by the Bill and Melinda Gates Foundation (BMGF) (INV-047051). The funders has no role on the design and interpretation of the data and model formulation and analysis.

### Author Declarations

Routine data for Kebbi State from January 2015 - May 2024 was obtained from National Malaria Data Repository (NMDR) of NMEP. The datasets used in our study had been de-identified prior to use in this study.

## References

[1] WHO. Fact sheet about malaria. https://www.who.int/news-room/fact-sheets/detail/malaria. [Accessed 11-02-2025].

[2] Maureen Coetzee. Key to the females of afrotropical anopheles mosquitoes (diptera: Culicidae). Malaria journal, 19: 1–20, 2020.

[3] Nelly O Kusimo, David E Matthew, Blessing A Olasunkanmi, Matthew E Agwae, and Michael O Kusimo. Understanding the importance of malaria control tools by pregnant and nursing mothers is key to ending malaria burden in nigeria: A case study of eight communities in south-south nigeria. International NGO Journal, 14(5): 64–69, 2019.

[4] World Health Organization. World Malaria Report 2021. World Health Organization, 2021. URL https://www.who.int/teams/global-malaria-programme/reports/world-malaria-report-2021.

[5] World Health Organization (WHO) Africa. Protecting families against malaria using long-lasting insecticide nets, n.d. URL https://www.afro.who.int/countries/nigeria/news/protecting-families-against-malaria-using-long-lasting-insecticide-nets. Accessed: 2025-02-16.

[6] National Malaria Elimination Programme (NMEP). Nigeria malaria indicator survey 2021 final report, 2021.

[7] World Health Organization. World malaria report 2023. World Health Organization, 2023.

[8] Federal ministry of health. Federal ministry of health, national malaria elimination programme (nmep), 2022. URL https://www.health.gov.ng/index.phpption=com_content&view=article&id=328&Itemid=550.

[9] National Malaria Elimination Programme (NMEP). Nigeria malaria strategic plan 2021-2025, 2021. URL https://nmcp.gov.ng/. Accessed: 2025-02-10.

[10] Wahab Adegbenro, Ebiwunmi T Oni, Ola Oba-Ado, Waheed A Folayan, Mr Olugboja Olafimihan, Tolu Arowolo, and Bamgboye M Afolabi. Replacement campaign of long lasting insecticide treated nets in ondo state, southwest nigeria, heartland of africa’s most efficient vector species. Diversity and Equality in Health and Care, 15(3): 95–103, 2018.

[11] Frank O Richards, Emmanuel Emukah, Patricia M Graves, Omeni Nkwocha, Lawrence Nwankwo, Lindsay Rakers, Aryc Mosher, Amy Patterson, Masayo Ozaki, Bertram EB Nwoke, et al. Community-wide distribution of long-lasting insecticidal nets can halt transmission of lymphatic filariasis in southeastern nigeria. The American journal of tropical medicine and hygiene, 89(3): 578, 2013.

[12] Ishaq Aliyu Abdulkarim, Ismaila Ibrahim Yakudima, Jamila Garba Abdullahi, and Yusuf M Adamu. Geographical analysis of malaria in nigeria–spatiotemporal patterns of national and subnational incidence. In Health and Medical Geography in Africa: Methods, Applications and Development Linkages, pages 185–209. Springer, 2023.

[13] Jash Mehta. Optimizing malaria control in nigeria: A comprehensive review of llin effectiveness and policy frameworks, 2024.

[14] Achuyt Bhattarai, Abdullah S Ali, S Patrick Kachur, Andreas Mårtensson, Ali K Abbas, Rashid Khatib, Abdul-wahiyd Al-Mafazy, Mahdi Ramsan, Guida Rotllant, Jan F Gerstenmaier, et al. Impact of artemisinin-based combination therapy and insecticide-treated nets on malaria burden in zanzibar. PLoS medicine, 4(11):e309, 2007.

[15] Mafalda Viana, Angela Hughes, Jason Matthiopoulos, Hilary Ranson, and Heather M Ferguson. Delayed mortality effects cut the malaria transmission potential of insecticide-resistant mosquitoes. Proceedings of the National Academy of Sciences, 113(32): 8975–8980, 2016.

[16] Jalal-Eddeen Abubakar Saleh, Abdullahi Saddiq, and Akubue Augustine Uchenna. LLIN ownership, utilization, and malaria prevalence: An outlook at the 2015 Nigeria malaria indicator survey. Open Access Library Journal, 5(1): 1–3, 2018.

[17] Lucy C Okell, Chris J Drakeley, Teun Bousema, Christopher J M Whitty, and Azra C Ghani. Modelling the impact of artemisinin combination therapy and long-acting treatments on malaria transmission intensity. PLoS medicine, 5(11):e226, 2008.

[18] World Health Organization. Mathematical Modelling to Support Malaria Control and Elimination. https://iris.who.int/bitstream/handle/10665/87051/9789241500418_eng.pdf?sequence=1, 2010. [Accessed 17-02-2025].

[19] EA Bakare, BO Onasanya, S Hoskova-Mayerova, and O Olubosede. Analysis of control interventions against malaria in communities with limited resources. Analele ştiinţifice ale Universităţii” Ovidius” Constanţa. Seria Matematică, 29(2): 71–91, 2021.

[20] B Gimba and SI Bala. Modeling the impact of bed-net use and treatment on malaria transmission dynamics, int. Sch. Res. Notices, 6182492, 2017.

[21] Abdulaziz YA Mukhtar, Justin B Munyakazi, Rachid Ouifki, and Allan E Clark. Modelling the effect of bednet coverage on malaria transmission in south sudan. PLoS One, 13(6): e0198280, 2018.

[22] Calistus N Ngonghala, Sara Y Del Valle, Ruijun Zhao, and Jemal Mohammed-Awel. Quantifying the impact of decay in bed-net efficacy on malaria transmission. Journal of theoretical biology, 363: 247–261, 2014.

[23] Calistus N Ngonghala. Assessing the impact of insecticide-treated nets in the face of insecticide resistance on malaria control. Journal of Theoretical Biology, 555:111281, 2022.

[24] Umar Boku IDRIS. Assessment of the implementation of demographic aspect of the nigeria’s national population policy in kebbi state, nigeria. 2023.

[25] Seyni Mamoudou. Economic and political factor of songhay empire the emergence of kebbi kingdom nigeria, c. 1500s. Open Journal of Social Sciences, 9(4): 332–345, 2021.

[26] S Wali, M Gada, I Hamisu, KJ Umar, and IG Abor. Evaluation of shallow groundwater in rural kebbi state, nw nigeria, using multivariate analysis: Implication for grou ndwater quality management. MOJ Eco Environ Sci, 7(3): 65–75, 2022.

[27] Usman Zayyanu Magawata and Abubakar Aliero Yahaya. Trends and variations of monthly solar radiation, temperature and rainfall data over birnin kebbi metropolis for the period of 2014-2016. Journal of Geography, Environment and Earth Science International, 21(2): 1–10, 2019.

[28] National Malaria Data Repository. National malaria data repository. https://nmdrnigeria.ng/dhis-web-commons/security/login.action, 2025. Contact. Accessed: 2025-05-22.

[29] Wilson L Mandala, Visopo Harawa, Fraction Dzinjalamala, and Dumizulu Tembo. The role of different components of the immune system against plasmodium falciparum malaria: Possible contribution towards malaria vaccine development. Molecular and biochemical parasitology, 246:111425, 2021.

[30] Emmanuel A Bakare, Elisha Bayode Are, Olusola E Abolarin, Samuel Ayodeji Osanyinlusi, Benitho Ngwu, and Obiaderi N Ubaka. Mathematical modelling and analysis of transmission dynamics of lassa fever. Journal of Applied Mathematics, 2020(1): 6131708, 2020.

[31] Kim A Lindblade, Laura Steinhardt, Aaron Samuels, S Patrick Kachur, and Laurence Slutsker. The silent threat: asymptomatic parasitemia and malaria transmission. Expert review of anti-infective therapy, 11(6): 623–639, 2013.

[32] Gholmreza Hassanpour, Mehdi Mohebali, Hojjat Zeraati, Ahmad Raeisi, and Hossein Keshavarz. Asymptomatic malaria and its challenges in the malaria elimination program in iran: a systematic review. Journal of Arthropod-Borne Diseases, 11(2): 172, 2017.

[33] Sandip Mandal, Somdatta Sinha, and Ram Rup Sarkar. A realistic host-vector transmission model for describing malaria prevalence pattern. Bulletin of mathematical biology, 75 (12):2499–2528, 2013.

[34] National population commission. nigeria demographic health survey (dhs). abuja, nigeria, 2018. URL https://www.dhsprogram.com/pubs/pdf/MIS20.pdf.

[35] CDC. About malaria, 2024. URL https://www.cdc.gov/malaria/about/index.html. Accessed: 2024-09-28.

[36] N. Chitnis, J. M. Hyman, and J. M. Cushing. Determining important parameters in the spread of malaria through the sensitivity analysis of a mathematical model. Bulletin of Mathematical Biology, 70: 1272–1296, 2008.

[37] WHO. Guidelines for laboratory and field testing of long-lasting insecticidal mosquito nets, 2005.

[38] Emmanuel Obi, Festus Okoh, Sean Blaufuss, Bolanle Olapeju, Joel Akilah, Okefu Oyale Okoko, Abidemi Okechukwu, Mark Maire, Kehinda Popoola, Muhammad Abdullahi Yahaya, et al. Monitoring the physical and insecticidal durability of the long-lasting insecticidal net dawaplus® 2.0 in three states in Nigeria. Malaria journal, 19: 1–19, 2020.

[39] Olusola Ojurongbe, Olubunmi A Lawal, Oyindamola O Abiodun, John A Okeniyi, Ayobami J Oyeniyi, and Oyeku A Oyelami. Efficacy of artemisinin combination therapy for the treatment of uncomplicated falciparum malaria in nigerian children. The journal of infection in developing Countries, 7(12): 975–982, 2013.

[40] David R Legates and Gregory J McCabe Jr. Evaluating the use of “goodness-of-fit” measures in hydrologic and hydroclimatic model validation. Water resources research, 35(1): 233–241, 1999.

[41] Cort J. Willmott. On the validation of models. Physical Geography, 2(2): 184–194, 1981. doi: 10.1080/02723646.1981.10642213.

[42] R. Kumar, M. Farzeen, A. Hafeez, B. K. Achakzai, M. Vankwani, M. Lal, and R. Somrongthong. Effectiveness of a health education intervention on the use of long-lasting insecticidal nets for the prevention of malaria in pregnant women of pakistan: a quasi-experimental study. Malaria Journal, 19: 1–10, 2020.

[43] M. K. Wubishet, G. Berhe, A. Adissu, and M. S. Tafa. Effectiveness of long-lasting insecticidal nets in prevention of malaria among individuals visiting health centres in ziway-dugda district, ethiopia: matched case–control study. Malaria Journal, 20(1): 301, 2021.

[44] UNICEF. Fighting malaria with long-lasting insecticidal nets (llins), 2022.

[45] J. F. Mosha, M. A. Kulkarni, E. Lukole, N. S. Matowo, C. Pitt, L. A. Messenger, and N. Protopopoff. Effectiveness and cost-effectiveness against malaria of three types of dual-active-ingredient long-lasting insecticidal nets (llins) compared with pyrethroid-only llins in tanzania: a four-arm, cluster-randomised trial. The Lancet, 399(10331): 1227–1241, 2022.

[46] World Health Organization. Who recommendations for achieving universal coverage with long-lasting insecticidal nets in malaria control, 2017. URL https://www.who.int/docs/default-source/malaria/mpac-documentation/mpac-oct2017-draft-updated-recommendations-universal-llin-coverage-session9.pdf.

[47] Pauline van den Driessche and James Watmough. Reproduction numbers and sub-threshold endemic equilibria for compartmental models of disease transmission. Mathematical Biosciences, 180(1–2): 29–48, 2002.

[48] Jeong-Heon Kim, Wei Su, and Yoon J Song. On stability of a polynomial. Journal of applied mathematics & informatics, 36(34):231–236, 2018.

[49] Bruce Elwyn Meserve. Fundamental concepts of algebra. Courier Corporation, 1982.

